# Highway paving increased dengue transmission in the Peruvian Amazon

**DOI:** 10.1101/2024.11.15.24317406

**Authors:** Alyson L. Singleton, Andres G. Lescano, Andrew J. MacDonald, Lisa Mandle, Terrell J. Sipin, Kevin S. Martel, César V. Munayco, Esteban D. R. Carrera, Gustavo A. Choque, Ana S. M. Bautista, Stephen P. Luby, Erin A. Mordecai

## Abstract

Human mobility drives the spread of many infectious diseases, yet the health impacts of mobility changes from new infrastructure development remain poorly understood and currently not accounted for in impact assessments. We present a novel application of a quasi-experimental approach to link mobility and infectious disease, leveraging historical road upgrades as a proxy for regional mobility changes. The 2009 paving of the Interoceanic Highway through the previously isolated Madre de Dios region of the Peruvian Amazon offered a natural experiment to evaluate how highway paving influences transmission of dengue virus, a high-burden mosquito-borne disease. We compared dengue incidence data from healthcare facilities near versus far from the highway before and after paving, while controlling for observable and unobservable confounding variables (a difference-in-differences causal inference approach). We found that the paving caused an additional 10,950 (95% CI: 3,186–18,715) dengue cases since 2009, a 403% (117–689%) increase in incidence rates in facilities near the highway in the 14 years since paving (2009–2022), compared to their pre-paving incidence rates. Our findings demonstrate the impact that infrastructure can have on dengue transmission, likely via its effects on human mobility. As a result, we advocate for future road construction plans in tropical regions to account for potential increases in dengue transmission during impact assessments.

## Introduction

Meeting United Nations Sustainable Development goals requires massive infrastructure build-out, especially in developing countries^1^. While there are many benefits promised by these infrastructure projects—such as increased access to labor opportunities, creation of water resources, and potential for economic growth—there are also many costs that have not been accounted for historically, such as harms to local environmental and human health^2,3^. Well-designed roads are critical for the distribution of resources and opportunities, and their growth is widespread; as of 2020, 12,263 km of road construction was planned in the Amazon alone^4^. Despite their importance to local development, roads drive changes in human mobility and migration that have been previously associated with increases in infectious disease transmission^5,6^. However, these effects remain a challenge to identify and are often not included in even the most comprehensive road assessment analyses^4^, likely due to the lack of causal analyses that disentangle disease from complex social-environmental processes. Current methods linking mobility and infectious disease also typically rely on cell phone or survey-based mobility data, which can be expensive, inaccessible, and/or biased due to lack of representativeness or recall bias^7,8^. To bridge these gaps, we present a novel application of a quasi-experimental approach where we suggest a causal link between highway paving and dengue virus transmission, leveraging the rapid paving of a highway through the Peruvian Amazon as a proxy for a discrete and significant increase in regional human movement and connectivity.

The Madre de Dios region of Peru is an ideal case study to investigate the relationships between human mobility and infectious disease given its recent infrastructure development, history of vector-borne disease, and high environmental suitability for vectors^9,10^. Madre de Dios is an 85,000 km^2^ department in the southwest Amazon in the southeast of Peru. In this region, most human population centers are distributed along rivers, secondary unpaved roads, and the only major paved road, the Interoceanic Highway, a controversial construction project completed in 2009 that intensified regional in-migration (e.g., people relocating from other parts of the country)^11^ and legal and illegal development^12^ (**Figure 1**). Touted as a project that would have socio-economic benefits for rural Peruvian communities, there have thus far been limited reports of economic or trade-based benefits in the area^13,14^. However, the transformation of an unpaved road into a paved highway spurred poverty-driven environmental degradation, such as accelerated deforestation and illegal gold mining^12^. There has been limited quantification of the health impacts of paving this section of highway; studies to date have been qualitative or correlational analyses^14,15^. Causal analyses of how highway construction may have influenced human health could help to build the evidence base on impacts of infrastructure projects in the Amazon.

**Figure 1:**
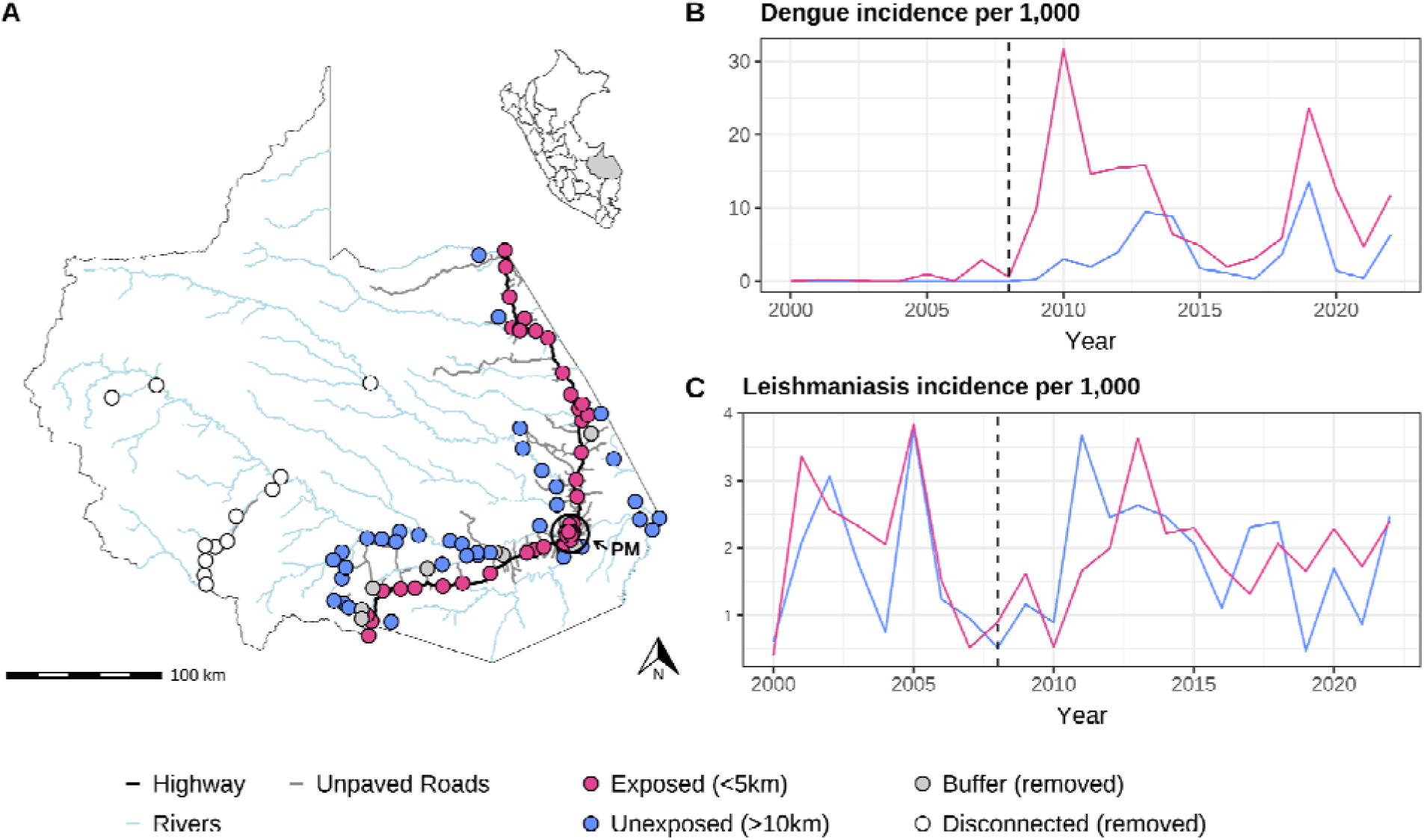
A map of Madre de Dios (A) displaying the Interoceanic highway (black line), unpaved roads (grey lines), rivers (blue lines), and the locations of all healthcare facilities colored by their exposure group (points). The region’s largest city, Puerto Maldonado, is indicated by the black circle and “PM” label. A more detailed map of Puerto Maldonado can be found in **Supplementary Figure S7**. Facilities within 5km of the road are considered exposed (pink), those >10km are considered unexposed (blue), and those between 5–10km from the road were excluded from analyses as a buffer against spillover effects of exposure to the highway (grey). Facilities disconnected from the region’s main transportation network were also excluded (white). Average annual dengue (B) and leishmaniasis (C) yearly incidence (cases per 1,000 per year) over the time period of interest colored by exposure group (buffer facilities are not included). Vertical dotted line at 2008 represents the last year before the highway was paved, which is the baseline year in our analyses.

We hypothesized that the highway paving most immediately affected dengue transmission through increases in mobility and connectivity both into and within the region, due to dengue’s specific epidemiology and ecology. Dengue virus is transmitted to humans by biting mosquitoes^16^. It is the most rapidly spreading mosquito-borne disease worldwide^17^ and has caused multiple record-breaking outbreaks over the last two years across the globe^18^. Qualitative analyses have reported that Madre de Dios community members believe that the highway construction facilitated increased dengue transmission^14^. Dengue was present throughout Madre de Dios well before the paving in 2009^10^ (**Figure 1B, Supplementary Figure S1A**), and its primary vector—*Aedes aegypti*—inhabits urban and peri-urban areas that line highway corridors^19^. Dengue is a highly localized disease, appearing in clusters at the household and neighborhood level^10^. Previous studies have established that diffusion of dengue through regional scales relies on human movement and migration^20^, given that its vector typically does not fly more than 1km^21^. We hypothesized that when the construction of the Interoceanic Highway further connected Madre de Dios to the bordering dengue-endemic regions of Acre, Brazil and Pando, Bolivia, increased human movement along the highway amplified virus importation and circulation (**Supplementary Figure S2**).

If increased human mobility is the main mechanism by which road paving affects infectious disease, then diseases that are less linked to human mobility should not show marked changes before and after paving in areas near the highway. In contrast to dengue, leishmaniasis—a parasitic disease caused by infection with *Leishmania* parasites transmitted by sandflies—was also well established in the region prior to road paving (**Figure 1C, Supplementary Figure S1B**) but is not expected to be tightly linked to human mobility and, by extension, to road paving^22,23^. Although sandflies have similar flight dispersal limitations^24^, they primarily occur in rural, forested, and frontier ecosystems farther from highway corridors^23^. In Madre de Dios, multiple *Leishmania* and sandfly species have been documented. Fourteen sandfly species have been found naturally infected with *Leishmania* spp., including *L. guyanensis* and *L. braziliensis*^23^. The highest infection rates were observed in *Lutzomyia sherlocki*, *Lu. walkeri*, and *Lu. llanosmartinsi*, with greater numbers of positive pools detected in communities farther from the highway^23^. Unlike *Aedes*, sandflies become infected by feeding on infectious animal reservoir hosts rather than humans (humans are dead-end hosts^25^). Therefore, while regional leishmaniasis transmission might have increased somewhat due to highway-driven land conversion, its eco-epidemiology minimizes the potential for it to be impacted by human movement along the highway.

We use leishmaniasis as a ‘negative control’ when testing impacts of the paving, in that we do not expect it to respond immediately to highway construction after controlling for land-use changes, but it might capture any impacts of highway paving on diagnostic capacity or healthcare seeking behavior. Notably, some evidence suggests that leishmaniasis in the Amazon may be more likely to manifest in immunologically naïve individuals^26^, suggesting that road-induced migration could increase susceptibility. However, we do not observe a post-paving shift in leishmaniasis trends (**Figure 1C**), supporting its use as a control for non-specific changes in healthcare access.

To establish whether the paving of the Interoceanic Highway in Madre de Dios caused an increase in dengue incidence rates, we conducted a difference-in-differences panel regression^27^, comparing dengue incidence rates in healthcare facilities (*n* = 81) within 5km of the newly paved highway (‘exposed’) and those at least 10km away from the highway (‘unexposed’), before and after the highway was paved in 2009 (**Figure 1**). This is a causal inference framework that approximates a before-after treatment-control experimental design and incorporates (1) fixed effects, which control for unobserved confounders (e.g., baseline dengue burden, large-scale political changes), and (2) observed control variables, which control for known dengue drivers (i.e., climate and land-use changes). We also tested whether or not the paving similarly affected leishmaniasis incidence rates, with the hypothesis that there was no impact on leishmaniasis because it is not directly affected by human mobility.

## Results

We found that the paving of the highway increased dengue incidence rates by 6.91 cases per 1,000 (95% CI: 2.01–11.81) in facilities within 5km of the highway compared to those outside 10km, after incorporating fixed effects and accounting for potentially confounding spatial, temporal, and environmental variation (**Table 1**, **Figure 2A**). This corresponded to a 403% (95% CI: 117–689%) increase in dengue incidence rates in healthcare facilities near the highway compared to their pre-paving incidence rates. Average yearly increases in exposed facilities ranged from 0.705 to 24.9 cases per 1,000 across the 14-year study period (**Figure 2A**, **Supplementary Table S1**). Yearly differences first peak two years after paving (2010), followed by a region-wide inter-epidemic period with minimal differences between groups (2014–2018), before a second peak emerged in 2019. Overall, we attributed 10,950 (95% CI: 3,186–18,715) dengue cases to the paving of the highway, accounting for 57.2% (95% CI: 16.6%–97.8%) of all dengue cases recorded in the region post highway construction (2009–2022). There was no systematic difference in trends in dengue incidence between healthcare facilities near the highway and those far from the highway before paving (coefficients remain indistinguishable from zero prior to 2008, **Figure 2A**), supporting the parallel trends assumption and implying that far facilities represent reasonable controls (i.e., counterfactuals) for facilities near the road.

**Figure 2:**
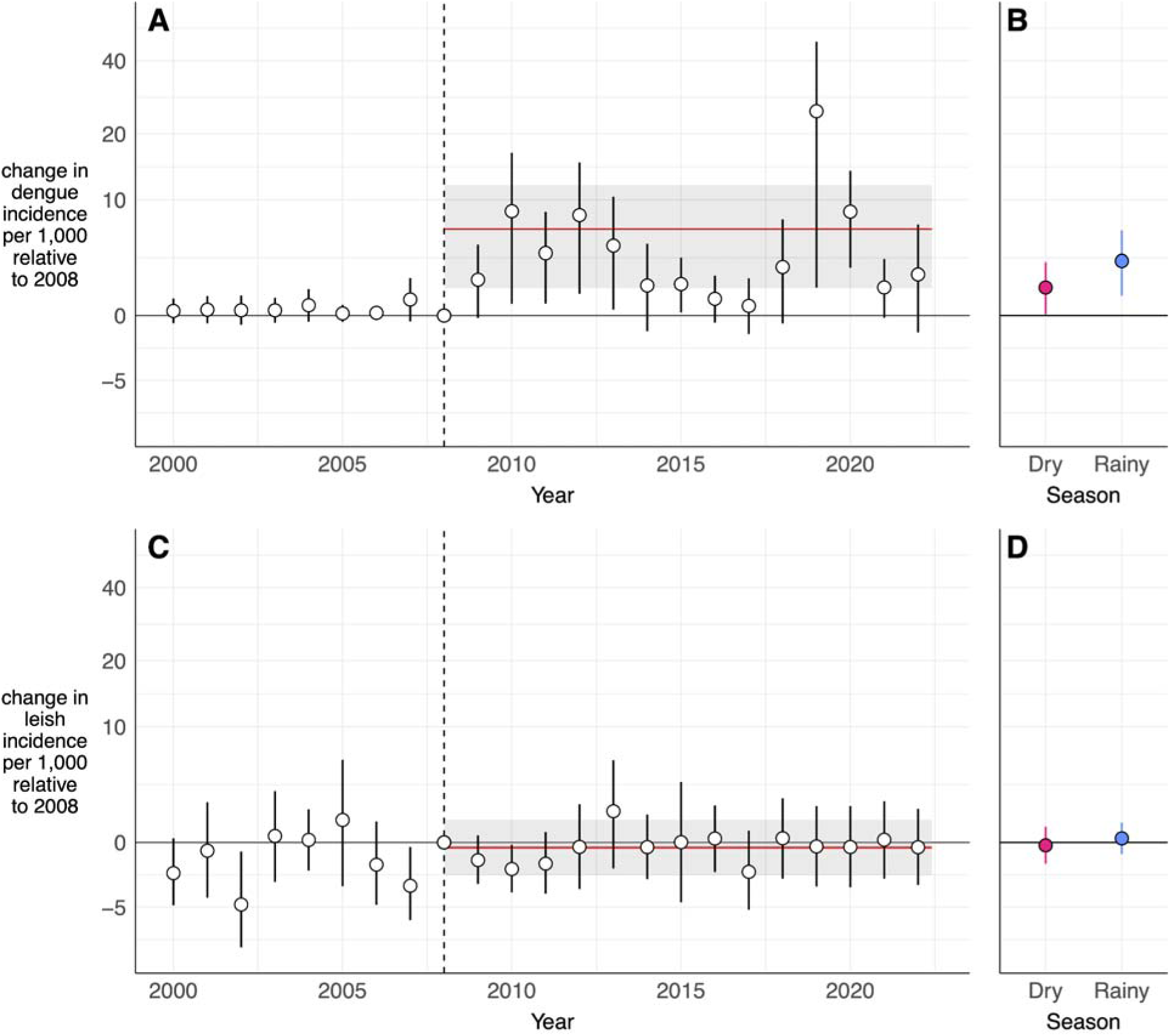
Dengue incidence increased substantially while leishmaniasis incidence remained stable in healthcare facilities near the highway compared to those far from the highway post-paving. Panels A and C display yearly estimation results from Equation 1 for dengue and leishmaniasis, respectively (treatment effects shown by points, 95% confidence intervals by error bars) and long-difference estimation results from Equation 2 (average treatment effect shown by the red line, 95% confidence interval by grey shading). Panels B and D display results for the biannual models aggregated across the full time period from Equation 2 for dengue and leishmaniasis, respectively. Note that the y-axis uses a log scale. The vertical dotted line at 2008 represents the last year before the highway was paved (baseline year). A table displaying yearly results (**Supplementary Table S1**) and a figure displaying disaggregated biannual estimates (**Supplementary Figure S10**) can be found in the **Supplementary Information**.

**Table 1:**
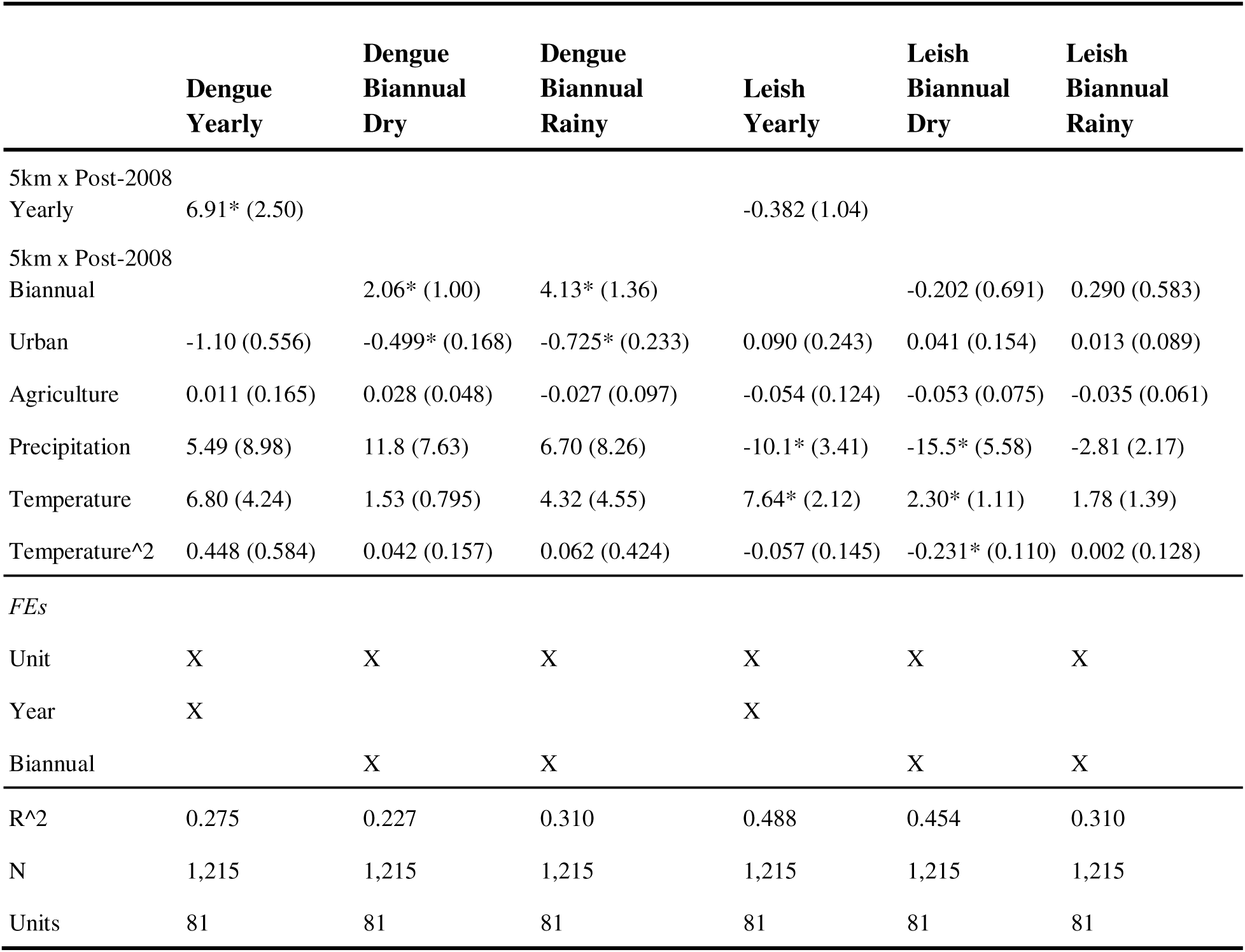
Dengue and leishmaniasis incidence long-differences estimation results. Estimation results from Equation 2 for yearly and biannual models. Yearly models include unit and year fixed effects, and biannual models include unit and biannual fixed effects. Standard errors are clustered at the group-level for all models and are displayed in parentheses next to each estimated coefficient. P-values indicating a statistical significance level below 0.05 are designated with an asterisk (*). **Supplementary Table S5** reports fixed effect terms for the dengue and leishmaniasis yearly models. **Supplementary Table S3** displays results for models with quadratic climate covariates, finding no major differences. **Supplementary Tables S4** displays results for models run using *glmmTMB*, also finding no major differences.

These findings were robust to various model specifications (**Supplementary Tables S2–S3**, **Supplementary Figures S3–S4**) and produced p-values < 0.01 when compared to permuted samples in permutation inference analyses (**Supplementary Figure S5**). Increases in precipitation were associated with increased dengue incidence, aligning with biological knowledge of *Aedes* vector habitat (**Table 1**). Seasonal (biannual) models aggregated across the study period revealed larger differences in dengue incidence between healthcare facilities near and far from the highway during rainy seasons (4.13 cases per 1,000, [95% CI: 1.46–6.79]), when the majority of cases occur, as compared to dry seasons (2.06 cases per 1,000, [95% CI: 0.09–1.46]) (**Figure 2B**).

As a ‘negative control’ that we do not expect to be immediately affected by increased human mobility, we used the same approach to examine impacts on leishmaniasis incidence before and after road paving. We found that leishmaniasis incidence did not differ systematically between healthcare facilities near and far from the highway either before or after highway paving (**Figure 2C–D**, **Table 1, Supplementary Table S1**). This supports our hypothesis that the observed changes in dengue incidence near the highway after paving were not solely driven by changes in diagnostic capacity or healthcare seeking behavior (which would have also appeared in the leishmaniasis incidence data), but are likely linked to changing epidemiology of dengue itself.

To better understand (1) which communities are at heightened risk of increased dengue transmission due to highway paving and (2) the extent of potential spatial spillover effects of exposure, we compared groups of healthcare facilities at increasing distances from the highway—1km, 5km, 10km, 15km, and 20km bins—to a control group of healthcare facilities >20km from the highway (**Figure 3B**). Healthcare facilities in the 1km bin (<1km) had significantly higher dengue incidence than healthcare facilities >20km from the highway post paving. Facilities in the 5km bin (>1km and <5km) and 10km bin (>5km and <10km) had higher dengue incidence than those >20km from the highway, although the effects were not statistically significant (**Figure 3A**). This suggests that clusters within 1km are primarily driving the effect observed in the main specification, which defines “exposed” facilities as those within 5km of the highway. Differences attenuated to zero as distance increased and were not statistically significant beyond 1km from the highway.

**Figure 3:**
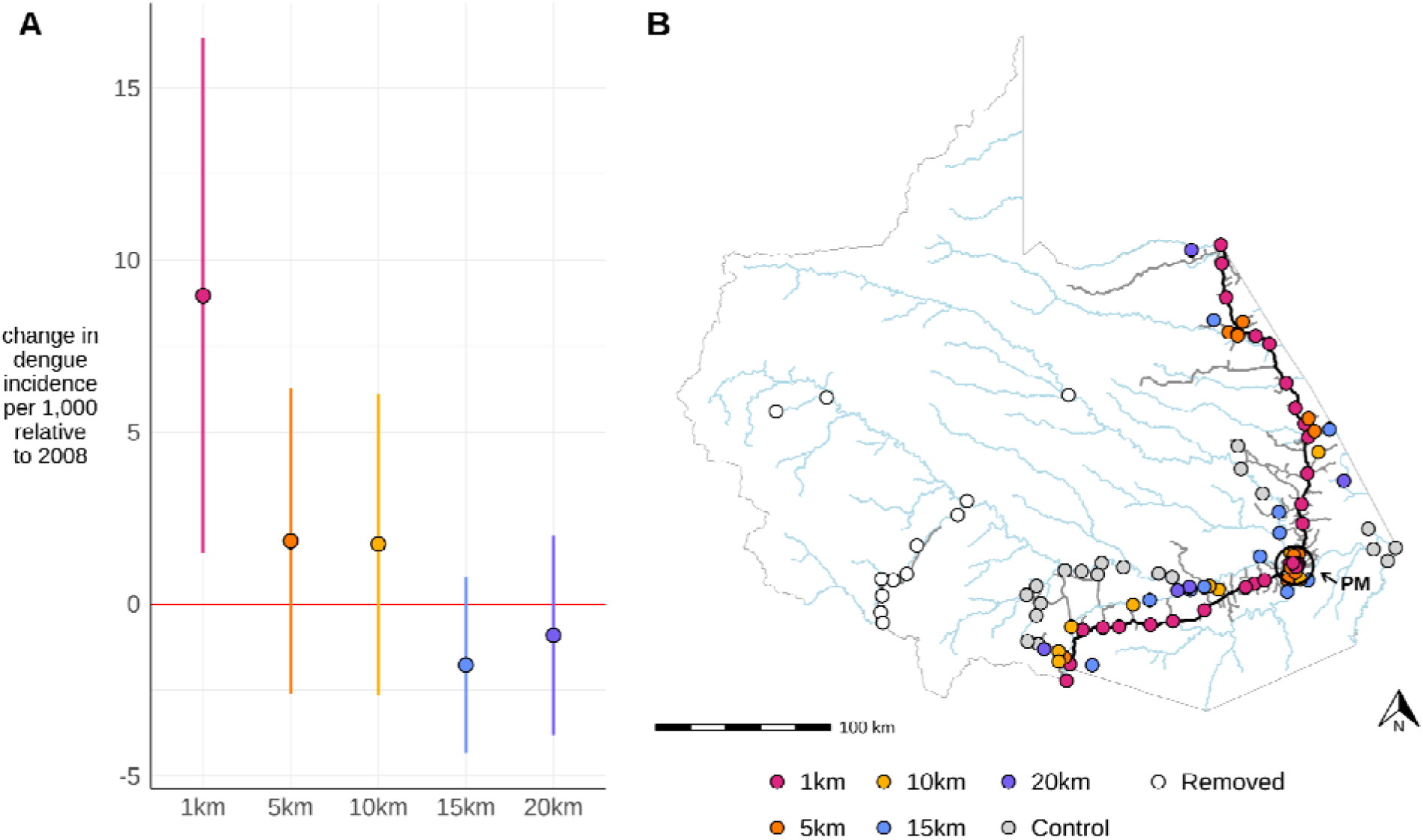
Effect of the highway attenuates when communities are at distances of greater than 1km from the highway. Panel A displays estimation results from Equation 2, where a model was run for each distance bin (colors, corresponding to Panel B). Average treatment effects are represented by points, and 95% bootstrapped confidence intervals are represented by error bars (bootstrapped standard errors clustered by spatial group). Panel B displays a map of the Madre de Dios region with healthcare facilities colored by their distance bin:1km (<1km, pink), 5km (>1km and <5km, orange), 10km (>5km and <10km, yellow), 15km (>10km and <15km, blue), 20km (>15km and <20km, purple), and control (>20km, gray). There were *n* = 33, *n* = 12, *n* = 9, *n* = 11, *n* = 5, and *n* = 20 facilities, respectively, in each distance bin. Facilities removed because they are disconnected from the region’s main transportation network are shown in white (*n* = 12). The map also displays the Interoceanic highway (black line), unpaved roads (grey lines), rivers (blue lines), and the region’s largest city, Puerto Maldonado, indicated by a black circle and “PM” label.

## Discussion

We found large and lasting impacts of the paving of the Interoceanic Highway on dengue incidence in a previously isolated region of the Peruvian Amazon: incidence rates increased by 6.91 cases per 1,000 (95% CI: 2.01–11.81) in healthcare facilities near versus far from the highway (**Table 1**, **Figure 2A**). While we do not directly test the mechanism, we hypothesize that the significant increase in dengue burden was driven by increased human mobility into and within the region (**Supplementary Figure S8**), leading to increased virus importation and circulation (**Supplementary Figure S2**). We extend previous research^5,6^ that associates highway construction with increased dengue transmission by offering causal estimates of the impact of road paving on dengue while controlling for potentially confounding variables. These estimates are robust to a suite of alternative model specifications (**Supplementary Table S2–S3, Supplementary Figures S3**–**S4**). We also conducted a ‘negative control’ analysis as a way to check for systematic changes that could confound our estimates, such as healthcare-seeking or case-reporting behaviors, and found that for leishmaniasis, a disease not expected to change with human mobility, there is no measurable effect of road paving on incidence (**Figure 2C–D**).

Our primary hypothesized mechanism for this effect is that highway paving facilitated regional human movement changes that caused additional dengue outbreaks to be seeded along the highway corridor. The highway increased connectivity to Brazil and Bolivia, both of which are dengue hyper-endemic, with regular co-circulation of multiple serotypes and genotypes^5,28^ (**Supplementary Figure S2**). Another relevant dynamic is the in-migration that occurred after the highway was built, with people moving to Madre de Dios from the nearby, newly connected Andes mountains and other non-endemic regions^11^ (**Supplementary Figure S6**). This increased the number of susceptible people living in the area. Although we directly account for yearly population changes when calculating incidence (**Supplementary Figure S6**) and determine that dengue cases increased more than what would have been expected just due to population growth, we do not explicitly include susceptibility dynamics in our model. However, if changes in susceptibility were a major driver, we might also expect to see increases in leishmaniasis, which can be more common among immunologically naïve individuals^26^. We detect no increase in leishmaniasis incidence following road paving.

Effects of highway paving on dengue were larger during rainy seasons as compared to dry, even after explicitly controlling for precipitation and including biannual fixed effects (**Figure 2B**). This could be due to the larger dengue burden during the rainy season enabling larger differences to occur between healthcare facilities near and far from the highway. Alternatively, this could be due to increased differences in mobility during the rainy season; unpaved roads often become impassable during the rainy season, while the highway is less affected by increased precipitation^29^. However, rivers can also become easier to navigate during the rainy season. Therefore, depending on whether remote communities rely more on unpaved roads or rivers, mobility may persist to a higher degree along the highway, potentially causing larger dengue differences between healthcare facilities near and far from the highway in the rainy season. Further supporting the role of movement and accessibility of the road, we found that communities within 1km from the highway experienced significantly increased dengue transmission after highway paving, while communities farther than 1km from the highway did not (**Figure 3**). This helps to define the spatial scale of impacts of highway development projects on dengue, with applications for designing control interventions.

Although this work isolates a plausible causal impact of highway paving on dengue transmission, our quasi-experimental approach does not allow us to definitively identify the specific mechanism that caused this effect. We hypothesize the main mechanism to be the increase in human movement along the corridor, supported by the fact that (1) we only see an immediate effect on dengue and not on leishmaniasis, (2) the effect is large even when explicitly controlling for resulting in-migration and land-use changes, and (3) the estimated effect attenuates at distances (>1km) less likely to be affected by mobility around the highway (and less likely to have vectors present, as discussed below). We include Peruvian governmental traffic volume data on interprovincial passenger traffic, estimated general vehicle fleet, national cargo vehicle fleet, and toll unit flow. These data show increases during the study period in the focal region but not in the demographically similar Amazonian region of Loreto, which did not experience road paving during this period (**Supplementary Figure S8**)^30^.

While changes in mobility and connectivity are parsimonious and plausible mechanisms behind the observed increases in dengue following highway paving, there are a few alternative phenomena that could have also contributed to the identified effect. Although we account for annual, facility-specific population changes, this does not allow us to isolate the impact of population growth on dengue transmission. While parts of the region were already environmentally suitable for dengue (**Supplementary Figure S1**), sustained transmission may have required both increased population density and greater virus circulation. Relatedly, we are also not able to include susceptibility dynamics in our model, which are known to be important for dengue due to antibody-dependent enhancement between dengue serotypes. However, it is important to note that large interannual changes in dengue outbreaks, for example following the introduction of a new serotype, are accounted for in the year fixed effects except to the extent that they differentially affect near versus far healthcare facilities. Lastly, highway construction practices more generally may have contributed to early dengue transmission by introducing susceptible workers and creating conditions favorable for *Aedes* breeding—patterns seen in other settings^31,32^. While we lack construction-specific data, results remain unchanged when the exposure year is shifted earlier, suggesting limited early effects prior to the paving project being completed (**Supplementary Figure S4**).

A key limitation of this study is the absence of vector presence or abundance data at the facility level. This restricts our ability to account for a critical condition for dengue transmission: the local presence of Aedes aegypti. Evidence from Loreto, the other heavily populated region of the Peruvian Amazon, shows that Ae. aegypti spreads along transportation corridors—rivers and highways—via human travel in boats and cars^19,33,34^. Vector density also appears to decline sharply with distance from rivers, suggesting that more remote communities in our study area may have had limited or no Aedes presence. This could bias our estimates downward: in facilities lacking vectors, reported dengue cases are likely imported or misdiagnosed, diluting observed effects by artificially increasing dengue incidence in the unexposed group. To partially address this concern, we exclude facilities that are disconnected from the region’s main transit network (**Figure 1A**, grey points) and conduct a robustness check that only retains facilities reporting at least one dengue case during the study period (**Supplementary Figure S3**, **Supplementary Table S2**). Additionally, although we control for urban and agricultural area changes, we were not able to control for hyper-localized habitat changes that might have implications for facility-level vector abundance and transmission capabilities, such as water storage and garbage practices that can influence breeding habitat.

Other limitations of this work include that different communities were exposed to their stretch of highway being paved at different times. Therefore, some units may have experienced exposure before 2009, resulting in an underestimation of the true effect size (although we find results to be robust to alternate choices of exposure year, **Supplementary Figure S4**).

Additionally, even after removing some units to create a ‘buffer’ against spatial spillover, some spillover likely remains throughout the unexposed units, also resulting in potential underestimation. Finally, although we attempt to control for development differences between exposed and unexposed groups through measuring yearly changes in urban area, it is possible that healthcare facilities near the highway developed increased capacity for diagnosing dengue compared to those far from the highway post paving. However, we show our results are robust to the inclusion of “probable” cases (**Supplementary Table S2, Supplementary Figure S3**), alleviating concerns of potential distortions due to the availability of test supplies. We also show that the number of facilities reporting at least one dengue or leishmaniasis case was comparable near and far from the highway both before and after the highway was paved (**Supplementary Figure S1**).

This work demonstrates the importance of including human health outcomes, such as infectious disease, in impact assessments when planning infrastructure development projects^3,35^. Our analysis establishes that highway construction through dengue-endemic areas makes communities uniquely vulnerable to dengue transmission. This work is especially timely because two additional highway construction projects through Madre de Dios were approved in 2023, over the protest of local communities and Indigenous groups^29,36^. Outside of the Amazon, other dengue-endemic tropical regions are also planning highway construction projects, including in Central America, Central Africa, and Southeast Asia^37^. Given the measurable and significant impact that road construction had on local dengue transmission in the Madre de Dios region, our findings contribute to understanding the social, environmental, health, and economic tradeoffs of this proposed infrastructure development. Should these infrastructure projects be approved, our work highlights the importance of coordinating with local, regional, and national healthcare officials at the outset of infrastructure project planning to forecast health impacts and target public health resources and interventions. Potential human health interventions could include targeted vector control strategies along road corridors^38^, investment in water and sanitation infrastructure to reduce mosquito breeding sites, and health system strengthening along highway corridors, such as increased staffing, priority access to new dengue vaccines once approved^39,40^, and deployment of dengue rapid diagnostic tests for early case detection ^41,42^.

We contribute to a growing body of literature that demonstrates the impact of infrastructure on human health, but dengue is only one of many infectious diseases that may be impacted by changes in human movement and development ^43–45^. Future work can help to identify the responses of other diseases to road building and other infrastructure development in the Amazon and across the globe. Generating this evidence will be key for guiding informed decision-making on meeting our development goals, while proactively mitigating any potential harms to local human and environmental health.

## Materials and methods

### Inclusion and ethics statement

The Stanford University IRB office determined that this analysis did not qualify as Human Subjects Research as defined in 45 CFR 46.102(e)(5), given that the data did not contain any identifiable information. All authors of this study have fulfilled the criteria for authorship through their participation in study design, study implementation, and/or data ownership. Roles and responsibilities were agreed upon among collaborators. This work includes findings that are locally relevant, which have been determined in collaboration with local partners. This research was not severely restricted or prohibited in the setting of the researchers, and does not result in stigmatization, incrimination, discrimination or personal risk to participants. Local and regional research relevant to our study was taken into account in citations.

### Data collection and preparation

De-identified, anonymized dengue and leishmaniasis case data reported from 2000–2022 were provided by the Regional Health Directorate of Madre de Dios (DIRESA^46^). Both dengue and leishmaniasis are reportable infectious diseases and are included in mandatory surveillance for each healthcare facility (*n* = 102) in Madre de Dios. They were the two most commonly reported infectious diseases in the region; no other diseases reported sufficient incidence to be included in this study. Case data included diagnosis code, week and year of diagnosis, method of diagnosis, healthcare facility name, and healthcare facility code. We identified latitude and longitude for each healthcare facility using a combination of government sources and the *ubigeo* package in R^47,48^.

Of 25,211 total dengue cases, 94.1% were laboratory confirmed and 5.9% were probable (see **Supplementary Information** for diagnosis methods). We included only confirmed cases in the main model and conducted a sensitivity analysis that included both confirmed and probable cases (**Supplementary Table S2**, **Supplementary Figure S3**). Of 15,507 total leishmaniasis cases, 99.4% of cases were laboratory confirmed and 0.6% were probable. We similarly included only confirmed leishmaniasis cases. 86.6% of leishmaniasis cases were cutaneous leishmaniasis and 13.4% were mucocutaneous.

We calculated facility-specific, yearly population measures by integrating healthcare facility population data from DIRESA and WorldPop^46,49^ (**Supplementary Figure S6A**, see **Supplementary Information**). Remotely-sensed environmental covariate data was downloaded in 5km^2^ circular buffer regions around each healthcare facility and included explicitly in models as control variables. First, we calculated the total yearly urban and agricultural area using Mapbiomas—a publicly available dataset of classified annual land-use maps of the Amazon basin with a spatial resolution of 30m^2^ (**Supplementary Figures S6B and S6C**^50^). Second, we calculated monthly mean temperature and accumulated precipitation using the ERA5 Land Monthly Aggregated dataset, which provides climate data at a spatial resolution of 11km^2^ ^51^. Monthly values were averaged or summed annually and biannually depending on each model’s temporal resolution.

To address any spatial autocorrelation occurring as a result of overlapping 5km^2^ buffer areas, we designated “spatial groups” of health facilities that were within 7.5km of one another (resulting in substantial buffer overlap). Clustering was done using an iterative hierarchical clustering approach that joined healthcare facilities in groups within the pre-specified distance of 7.5km (*hclust* and *cutree* functions from the *stats* R package). Facilities with minimal or no overlapping buffer areas were designated as their own spatial group. We clustered all modeled standard errors based on these spatial groupings (**Supplementary Figure S7**).

### Interoceanic Highway

We leveraged the paving of the highway as a natural experiment, or shock, hypothesized to result in a sudden increase in mobility-driven dengue transmission in areas near the highway. This paving is plausibly exogenous to our system (i.e., the decision to pave and location of the road paving did not depend on dengue incidence at baseline), given that the road was paved as a part of the larger Interoceanic Highway construction project, spanning from the Atlantic coast of Brazil to the Pacific coast of Peru^52^. The road paving site was selected to pass through the tri-border area at the northeast end of the region where Peru borders Brazil and Bolivia and then minimize costs to reach Puerto Maldonado, the region’s capital, and eventually Cuzco ^52^ (**Supplementary Figure S2**).

The construction of the Interoceanic Highway through Peru took place from 2006 to 2010^53^. Further investigation confirmed the section of highway specific to Madre de Dios (Tramo 3, Inambari–Iñapari) was completed in 2009^54^. While paving through Madre de Dios occurred from 2007 to 2009^54–56^, our goal was to capture when regional human connectivity and movement shifted regionally, rather than the moment households individually experienced paving adjacent to their community. Therefore, we designate 2009 as the year when exposure began for all spatial units and 2008 as our ‘baseline’ year (i.e., the last year before exposure began). As a robustness check, we also conduct a sensitivity analysis comparing models in which the exposure year was designated as either 2007 or 2008 (corresponding to baseline years of 2006 and 2007, respectively; **Supplementary Figure S4**). The paving of the highway shortened the time it takes to cross the region from a multi-day trip to five hours^57^. Supplementary Peruvian governmental data shows corresponding increases in traffic volumes in Madre de Dios after highway paving (**Supplementary Figure S8)**^30^, and community members report increased tourism, increased traffic, and faster transportation in stakeholder surveys^58^.

This study design designates healthcare facilities within 5km of the highway as ‘exposed’ (*n* = 45) and those further than 10km from the road as ‘unexposed’ (*n* = 36), using distance as a proxy for connectivity to the highway. The 5km cutoff represents a conservative definition of exposure, given the typical flight range of *Aedes* vectors is less than 1km^21^. We also conducted a supplementary analysis following methods from Carrasco-Escobar *et al.*, 2020^59^ to calculate the travel time from each healthcare facility to the highway, which shows agreement between the distance proxies and travel time (**Supplementary Table S4**, more methodological detail in **Supplementary Table S4** caption). Understanding that there is likely spatial spillover of exposure, we removed healthcare facilities between 5–10km from the highway as a ‘buffer’ (*n* = 9, **Figure 1A**). We tested the sensitivity of the model to both the boundary and buffer specifications (**Supplementary Table S2**, **Supplementary Figure S3**). Finally, we removed healthcare facilities that were substantially disconnected from the region’s main transportation network, making them less comparable controls (*n* = 12, **Figure 1A**).

The key identifying assumption in the difference-in-differences design is that both groups of healthcare facilities would have seen dengue incidence develop along parallel trends in the absence of the road paving. Visually, the parallel trends assumption appears defensible, with dengue and leishmaniasis incidence following parallel trajectories in facilities designated exposed versus unexposed prior to paving (i.e., pre-exposure; **Figures 1B and 1C**). Additionally, all remaining healthcare facilities far from the highway are connected to the region’s larger transportation network either by unpaved roads or rivers, making their connection to the region’s transportation network comparable to that of the healthcare facilities near the highway prior to exposure (**Figure 1A**).

### Statistical models

To identify the effect of paving the Interoceanic Highway on dengue transmission, we used a difference-in-differences panel regression^27^, comparing healthcare facilities within 5km of the newly paved highway (‘exposed’) and those outside 10km (‘unexposed’), before and after the highway was paved in 2009. The model takes the form (Equation 1):

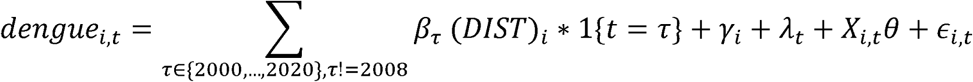

where *i* is unit (healthcare facilities) and *t* is year. Our outcome of interest, *dengue_i,t_*, is the dengue incidence per 1,000 in healthcare facility *i* for year *t*. Our exposure variable, DIST, is a dummy (binary) variable that equals one for exposed facilities (<5km) and zero for unexposed facilities(>10km). The DIST dummy variable interacts with year dummy variables (one for each year of interest and zero otherwise). We omit 2008, the year before the highway was paved, as the baseline year. The coefficients, β’s, on the interaction term then estimate the response in our outcome of interest following the road paving. Each coefficient estimates the difference in incidence rates between near and far healthcare facilities for each year in our set of τ’s, compared to 2008. We include unit (i.e., healthcare facility) fixed effects (FEs), γ*_i_*, to help account for any time-invariant healthcare facility characteristics, such as differences in their invasion history of *Aedes* mosquitoes, proximity to protected areas, and baseline dengue burden at the community level. FEs are a set of dummy variables that each represent a group (e.g., healthcare facilities), demeaning the data within each group. We also include year FEs, λ*_t_*, to account for large-scale changes that might impact dengue comparably across all healthcare facilities over time, such as El Niño events, economic recessions, and global pandemics. Finally, we include four observable controls, denoted by the matrix X*_i,t_*, to explicitly control for any variation due to changes in urban area, agricultural area, temperature, or precipitation. All controls are included as linear terms except for temperature, which is included as a quadratic function given its well-established non-linear relationship with vector-borne transmission potential.^60^ The θ vector recovers our covariate coefficients. Any remaining unobserved variation is captured by the error term, ε*_i,t_*.

If our hypothesis that the highway paving increased dengue transmission in healthcare facilities near the highway is correct, we expect the β coefficients will diverge from zero and become positive following 2008. Coefficients remaining at zero prior to 2008 would support the parallel trends assumption and the validity of using the unexposed group as controls. As a ‘negative control’ test, we conducted the same analysis with leishmaniasis incidence as our outcome, with the expectation of no significant differences in outcome between exposed and unexposed groups pre- or post-road paving.

Models were estimated using a Gaussian distribution with the *fixest*^61^ package in R and standard errors were clustered by the spatial groupings described above (see *Data collection and preparation*) to account for autocorrelation between pre- and post-road paving in the same healthcare facilities and facilities with overlapping environmental covariates. Although Poisson models are standard for count data, they assume multiplicative effects and perform poorly when many units transition from zero to non-zero counts. In contrast, Gaussian models estimate additive changes without requiring data transformation or exclusion of zeros^62^, making them better suited for this study setting where we observe dengue emergence in previously unaffected communities.

As robustness checks, we tested the sensitivity of model estimates to excluding Puerto Maldonado (the region’s capital), removing urban and agricultural area covariates, incorporating population weighting, changing the spatial boundary delineating exposed and unexposed groups, changing the buffer zone size between exposed and unexposed groups, including both confirmed and probable cases, and only including spatial units reporting at least one case of dengue during the study period (**Supplementary Table S2, Supplementary Figure S3**). We also compared a linear precipitation term to a quadratic form (**Supplementary Table S3**), changed the time of exposure to 2007 or 2008 (**Supplementary Figure S4**), and used the *glmmTMB*^63^ package instead of *fixest* (**Supplementary Table S6**). We report all *glmmTBM* derived estimates, including intercept terms in the **Supplementary Information** (**Supplementary Table S7**). Finally, we conducted permutation inference analysis, where we randomized our exposure variable using three methods—randomizing across facilities and years, randomizing across facilities while maintaining temporal structure, and randomizing across years while maintaining spatial structure—to establish that there are no spurious cross-sectional or temporal phenomena contributing to the observed effect (**Supplementary Figure S5**).

Because rainfall is highly seasonal in this region and dengue transmission peaks in the rainy season (**Supplementary Figure S9**), we also ran biannual models to account for heterogeneous effects across seasons (**Supplementary Figure S10**). These models were equivalently parameterized as in Equation 1 except *t* was biannual time periods (October–March, April–September for each year) that were constructed to align with the rainy season (months with above average monthly accumulated precipitation from 2000–2022, **Supplementary Figure S9**). Biannual models included biannual fixed effects in place of year fixed effects and used the dry season (April–September) of 2008 as the baseline.

Finally, we estimated aggregated versions of Equation 1 over all post-paving years to summarize average treatment effects and the total number of attributable cases (e.g., a long-differences model). We define a post-paving dummy variable equal to zero in 2008 and one from 2009 to 2022. The aggregated model takes the form (Equation 2):

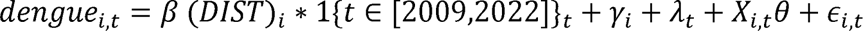

This is also the model specification used for aggregating biannual model average treatment effects (with biannual fixed effects substituted for year fixed effects, **Figure 2B** and **Figure 2D**). We also used the aggregated model to compare groups of healthcare facilities at increasing distances from the highway—1km (<1km), 5km (>1km and <5km), 10km (<5KM and >10km), 15km (>10km and <15km), and 20km (>15km and <20km) bins—to a control group of healthcare facilities >20km from the highway (**Figure 3A**). We calculated bootstrapped standard errors for this analysis given the small sample size of some of the distance bins (i.e., 1km, 5km, etc.). We bootstrapped by sampling healthcare facilities with replacement, retaining the full time series for each facility in the bootstrapped sample and maintaining the ratio of exposed to unexposed units for each bin. In all other analyses we used analytic standard errors.

### Percent change and attributable cases

We calculated percent changes in dengue incidence due to highway paving relative to the average dengue incidence rate in the exposed group directly prior to paving (i.e., 2007–2008), which was 1.71 dengue cases per 1,000 people. This baseline was chosen to represent the most conservative, pre-intervention risk level in what would become the “exposed” group. Percent changes in incidence were computed by dividing model estimates by this pre-exposure average. To estimate the number of cases attributable to highway paving, we calculated the expected number of dengue cases in the exposed group during the post-exposure period (2009–2022) by multiplying the pre-treatment incidence rate by the observed population in each year. We then multiplied the estimated percent increase by this expected number to derive the total number of excess dengue cases associated with highway paving, accounting for both fixed effects and time-varying covariates.

## Data availability

The disease case data used in this analysis was provided by the Peruvian Ministry of Health and is shared publicly at https://github.com/alyson-singleton/IOH_and_dengue. The repository also includes all population, environmental, traffic, and spatial data needed to reproduce the analysis.

## Code availability

All code and analysis for this study is available via https://github.com/alyson-singleton/IOH_and_dengue.

## Acknowledgements

We thank all the health personnel of the Madre de Dios region who contributed to generate this valuable data, as well as the Peruvian National Epidemiology Network (RENACE). We acknowledge and thank the members of the Mordecai and Luby Labs of Stanford University for their support and contribution to this work. We would also like to acknowledge computational resources from Google Cloud for Earth Engine. ALS was supported by the Jim and Gaye Pigott Fellowship through the Stanford Interdisciplinary Graduate Fellowship program at Stanford University. AGL is sponsored by Emerge, the Emerging Diseases Epidemiology Research Training grant D43 TW007393 awarded by the Fogarty International Center of the US National Institutes of Health. AJM acknowledges the USDA National Institute of Food and Agriculture (2023-68016-40683) and the National Science Foundation and Fogarty International Center (DEB-2011147; DEB-2339209). LM was supported by the National Science Foundation (DEB-2011147 with Fogarty International Center). EAM was supported by the National Science Foundation (DEB-2011147 with Fogarty International Center), the National Institutes of Health (R35GM133439, R01AI168097).

## Supplementary Methods

### Disease diagnosis

As defined by the Peruvian Ministry of Health, laboratory confirmed dengue cases are cases that produce a positive result from one or more of the following tests: “1) DENV isolation by cell culture, 2) qRT-PCR, 3) ELISA antigen NS1, 4) detection of IgM antibodies for dengue in a single sample by ELISA in dengue endemic areas, and/or 5) evidence of IgM seroconversion in paired samples, where the second sample should be taken after 14 days of the onset of symptoms in areas where there is no transmission of dengue (and these cases should have an epidemiological investigation)”^1^. Probable dengue cases are clinically diagnosed and must either meet case definition criteria set by the World Health Organization^2^ or have a direct epidemiological link to a confirmed case^1^.

Laboratory confirmed leishmaniasis cases are defined as cases producing a positive result from “1) a parasitological examination, 2) an immunological or histopathological clinical, and/or 3) a culture, whether or not in the form of a pruritic nodule.” Probable cases of cutaneous leishmaniasis were clinically diagnosed and defined as, “any person with progression to ulcerative or ulcerous-crusted lesions, shallow, rounded, non-painful, with well-defined borders and inflammatory signs, with evolution time of no less than four weeks and lack of response to conventional treatment. With a history of origin or residence in […] areas endemic to leishmaniasis.” Probable cases of mucocutaneous leishmaniasis were clinically diagnosed and defined as, “any person with a clinical picture characterized by raised granulomatous or ulcerative lesions of the nasal mucosa, mouth, white palate, pharynx, larynx or trachea, with a history of active or healed skin lesions, origin or residence in endemic areas^3,4^.” Our measure of leishmaniasis incidence combined laboratory confirmed leishmaniasis and mucocutaneous leishmaniasis cases, with a robustness check in which we include both types of probable cases in the analyses.

### Population calculation

Health facility population data was provided by DIRESA for the years 2009–2017 and 2020– 2022. This population data was linked to case data through the facility names in the disease case dataset. In years without DIRESA-provided population data, we imputed missing values with WorldPop data, a remotely-sensed population dataset that estimates the number of people living in each 100m pixel from 2000–2020.^7^ To align the two data sources, we scaled each facility-year WorldPop value by the average ratio between DIRESA and WorldPop values for that facility’s 5km^2^ buffer area, wherever both sources overlapped. We also anchored the imputed values to known DIRESA population counts in adjacent years and interpolated (two anchor points) or extrapolated (one anchor point) using the shape of WorldPop growth. This allowed us to preserve observed trajectories while maintaining consistency with the DIRESA-reported values. It is important to measure annual changes in each facility’s population due to the rapid in-migration and population growth in the area during the study period^8^ (**Supplementary Figure S5**). We use these population measures to derive incidence rates from disease case counts as a measure of changes in per-capita dengue risk, above and beyond changes in raw case counts that would be expected due to population growth alone.

## Supplementary Tables S1–S6

**Supplementary Table S1:**
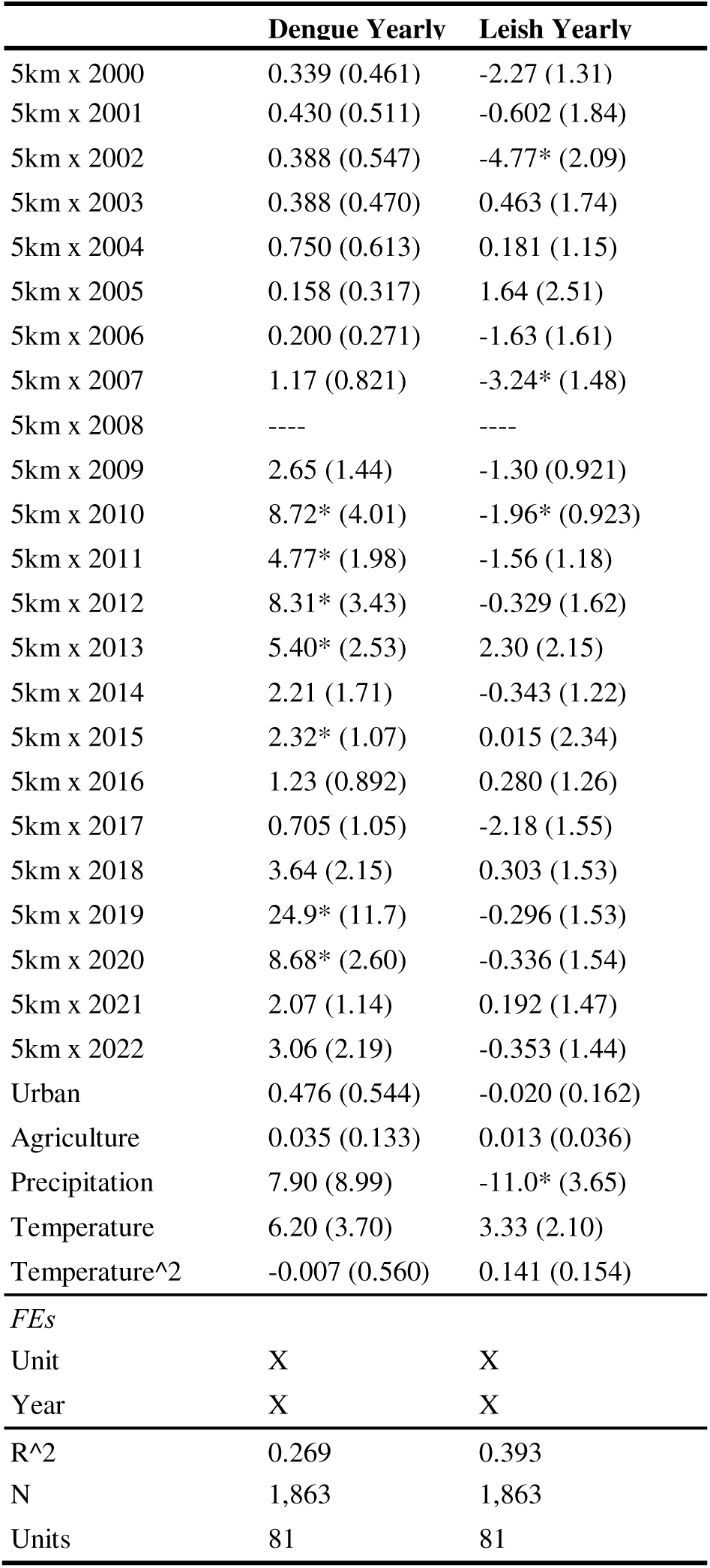
Dengue and leishmaniasis yearly estimation results (Equation 1). Standard errors are clustered by spatial group for all models and are displayed in parentheses next to each estimated coefficient. P-values indicating a statistical significance level below 0.05 are designated with an asterisk (*).

**Supplementary Table S2:** Dengue long-differences estimation results (Equation 2) are displayed for the following model specifications: main model, model excluding Puerto Maldonado (the region’s largest city), model excluding land-use covariates, population-weighted model, model designating <1KM from the highway as exposed and >1km from the highway as unexposed, model designating <10KM as exposed and >10km as unexposed, model with no spatial buffer (<5km exposed, >5km unexposed), model with a larger spatial buffer (<5km exposed, >20km unexposed), model including both confirmed and probable cases, and model including only spatial units reporting at least one dengue case during the study period. All models include unit and year fixed effects. Standard errors are clustered by spatial grouping for all models and are displayed in parentheses next to each estimated coefficient. P-values indicating a statistical significance level below 0.05 are designated with an asterisk (*). Corresponding yearly results are shown in **Supplementary Figure S3.**

|  | Main<br>model | No PM | No LUC | Pop<br>weight | 1km<br>boundary | 10km<br>boundary | No buffer<br>(<5km v<br>>5km) | Large<br>buffer<br>(<5km v<br>>20km) | Conf &<br>prob<br>cases | Units w<br>dengue |
| --- | --- | --- | --- | --- | --- | --- | --- | --- | --- | --- |
| 5km x Post-2008 | 6.91*<br>(2.50) | 4.07*<br>(1.49) | 6.02*<br>(1.78) | 12.1*<br>(4.14) |  |  | 6.27*<br>(2.47) | 7.09*<br>(2.65) | 7.16*<br>(2.48) | 7.44*<br>(2.70) |
| 1km x Post-2008 |  |  |  |  | 6.65*<br>(1.90) |  |  |  |  |  |
| 10km x<br>Post-2008 |  |  |  |  |  | 6.17*<br>(2.18) |  |  |  |  |
| Urban | -1.10<br>(0.556) | 1.46<br>(2.83) |  | -2.00*<br>(0.513) | -1.01*<br>(0.498) | -1.01*<br>(0.499) | -1.04*<br>(0.514) | -1.50*<br>(0.611) | -0.854<br>(0.576) | -1.49*<br>(0.729) |
| Agriculture | 0.011<br>(0.165) | 0.073<br>(0.158) |  | 0.060<br>(0.171) | 0.005<br>(0.150) | 0.020<br>(0.148) | 0.015<br>(0.150) | -0.144<br>(0.147) | 0.015<br>(0.167) | -0.014<br>(0.210) |
| Precipitation | 5.49<br>(8.98) | 0.800<br>(8.24) | 7.43<br>(9.31) | 4.35<br>(12.9) | 3.69<br>(7.10) | 3.61<br>(7.16) | 4.17<br>(7.27) | 8.10<br>(9.94) | 4.83<br>(9.59) | 1.88<br>(10.0) |
| Temperature | 6.80<br>(4.24) | 6.00<br>(4.12) | 7.00<br>(5.03) | 12.6*<br>(3.64) | 6.39<br>(3.79) | 6.71<br>(3.81) | 7.11<br>(3.90) | 7.50<br>(4.62) | 7.28<br>(4.98) | 5.39<br>(4.86) |
| Temperature<br>^2 | 0.448<br>(0.584) | 0.046<br>(0.508) | 0.210<br>(0.452) | 2.62*<br>(0.998) | 0.363<br>(0.580) | 0.376<br>(0.521) | 0.311<br>(0.542) | 0.478<br>(0.626) | 0.422<br>(0.606) | 0.525<br>(0.571) |
| R^2 | 0.275 | 0.232 | 0.273 | 0.412 | 0.280 | 0.280 | 0.280 | 0.277 | 0.277 | 0.288 |
| N | 1,215 | 1,065 | 1,215 | 1,215 | 1,350 | 1,350 | 1,350 | 1,050 | 1,215 | 915 |
| Units | 81 | 71 | 81 | 81 | 90 | 90 | 90 | 70 | 81 | 61 |

**Supplementary Table S3:** Dengue and leishmaniasis incidence long-differences estimation results (Equation 2) with the precipitation covariate as a quadratic function (rather than linear). Corresponds to Table 1 in main text. Yearly models include unit and year fixed effects, and biannual models include unit and biannual fixed effects. Standard errors are clustered by spatial grouping for all models and are displayed in parentheses next to each estimated coefficient. P-values indicating a statistical significance level below 0.05 are designated with an asterisk (*).

|  | Dengue Yearly | Dengue Biannual Dry | Dengue Biannual Rainy | Leish Yearly | Leish Biannual Dry | Leish Biannual Rainy |
| --- | --- | --- | --- | --- | --- | --- |
| 5km x Post-2008 Yearly | 6.98* (2.52) |  |  | -0.257 (1.05) |  |  |
| 5km x Post-2008 Biannual |  | 2.16* (1.05) | 4.15* (1.34) |  | -0.158 (0.692) | 0.305 (0.585) |
| Urban | -1.08 (0.551) | -0.469* (0.150) | -0.707* (0.232) | 0.111 (0.241) | 0.053 (0.152) | 0.025 (0.087) |
| Agriculture | 0.016 (0.166) | 0.033 (0.047) | -0.016 (0.098) | -0.044 (0.124) | -0.051 (0.074) | -0.028 (0.060) |
| Precipitation | 7.81 (10.6) | -2.24 (8.17) | 14.2 (17.5) | -5.80 (4.08) | -21.2* (8.31) | 2.13 (3.95) |
| Precipitation^2 | -5.56 (14.6) | -64.7 (50.5) | -11.3 (16.2) | -10.3 (6.52) | -26.4 (29.0) | -7.46 (4.67) |
| Temperature | 7.05 (4.51) | 2.07* (0.919) | 5.31 (5.54) | 8.09* (2.18) | 2.52* (1.12) | 2.42 (1.35) |
| Temperature^2 | 0.553 (0.731) | 0.090 (0.150) | 0.144 (0.430) | 0.137 (0.153) | -0.212* (0.105) | 0.056 (0.129) |
| *FEs* |  |  |  |  |  |  |
| Unit | X | X | X | X | X | X |
| Year | X |  |  | X |  |  |
| Biannual |  | X | X |  | X | X |
| R^2 | 0.275 | 0.228 | 0.31 | 0.489 | 0.454 | 0.311 |
| N | 1,215 | 1,215 | 1,215 | 1,215 | 1,215 | 1,215 |
| Units | 81 | 81 | 81 | 81 | 81 | 81 |

**Supplementary Table S4:** Cumulative cost mapping analysis shows agreement between distance proxies (km) and travel time estimates (min) from the paved road. To calculate these values we followed methods from Carrasco-Escobar *et al.*, 2020^9^, which estimate the minimum travel time from each healthcare facility to the highway (using their publicly available code). We constructed a friction surface that calculates travel speeds for each pixel based on access to road networks (paved vs. unpaved), rivers, and various land cover types (also incorporating their slope and elevation, which can impact walking speed). A cumulative cost algorithm is then applied to compute the minimum possible travel time in minutes from each pixel to the nearest point on the highway, the results of which are shown below.

| **Distance Boundary** | **Median Travel Time to Interoceanic Highway (min)** |
| --- | --- |
| 1km | 0 |
| 5km | 9 |
| 10km | 26 |
| 15km | 32 |
| 20km | 130 |
| 40km | 154 |
| >40km | 1139 |

**Supplementary Table S5:**
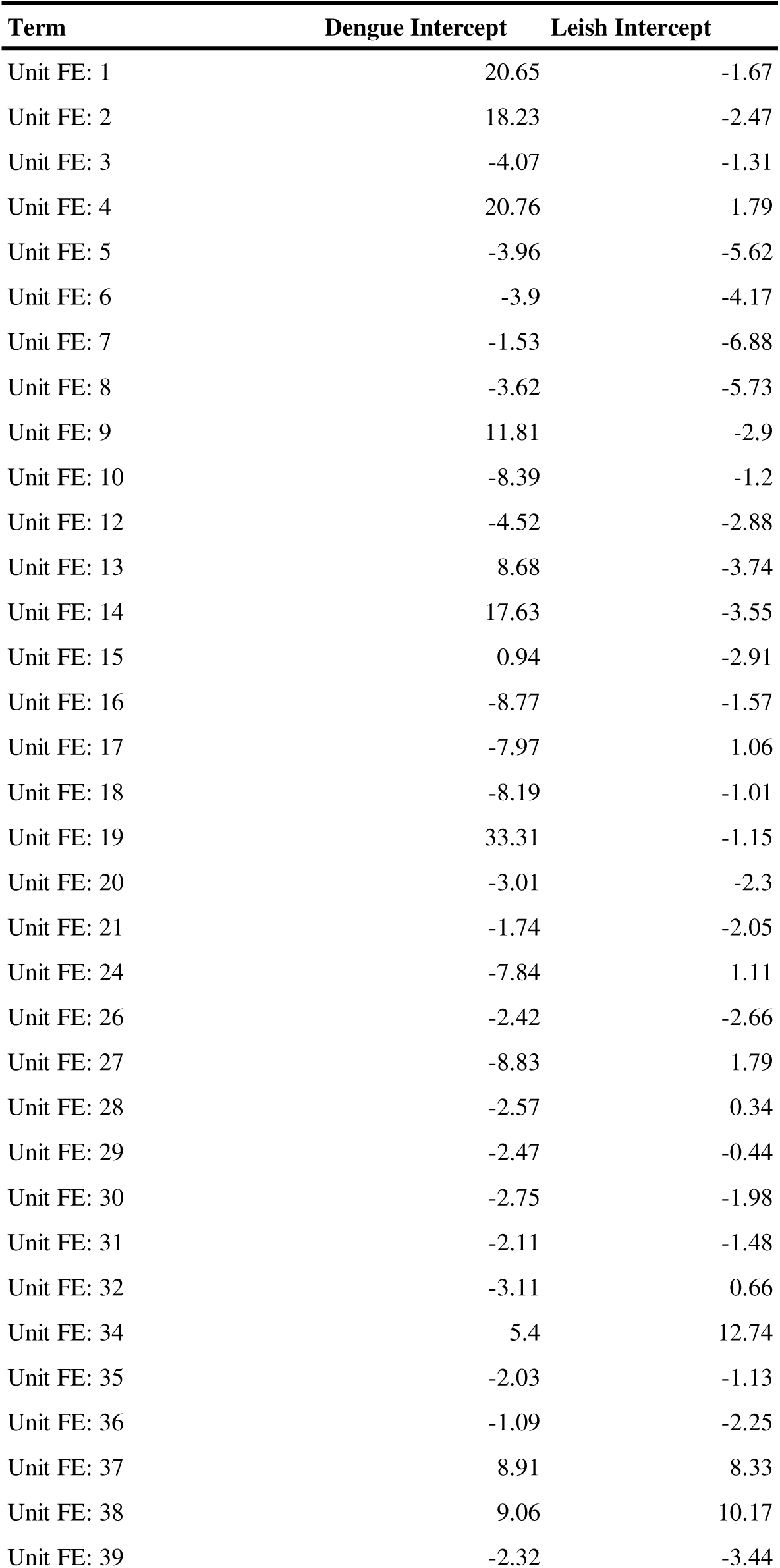
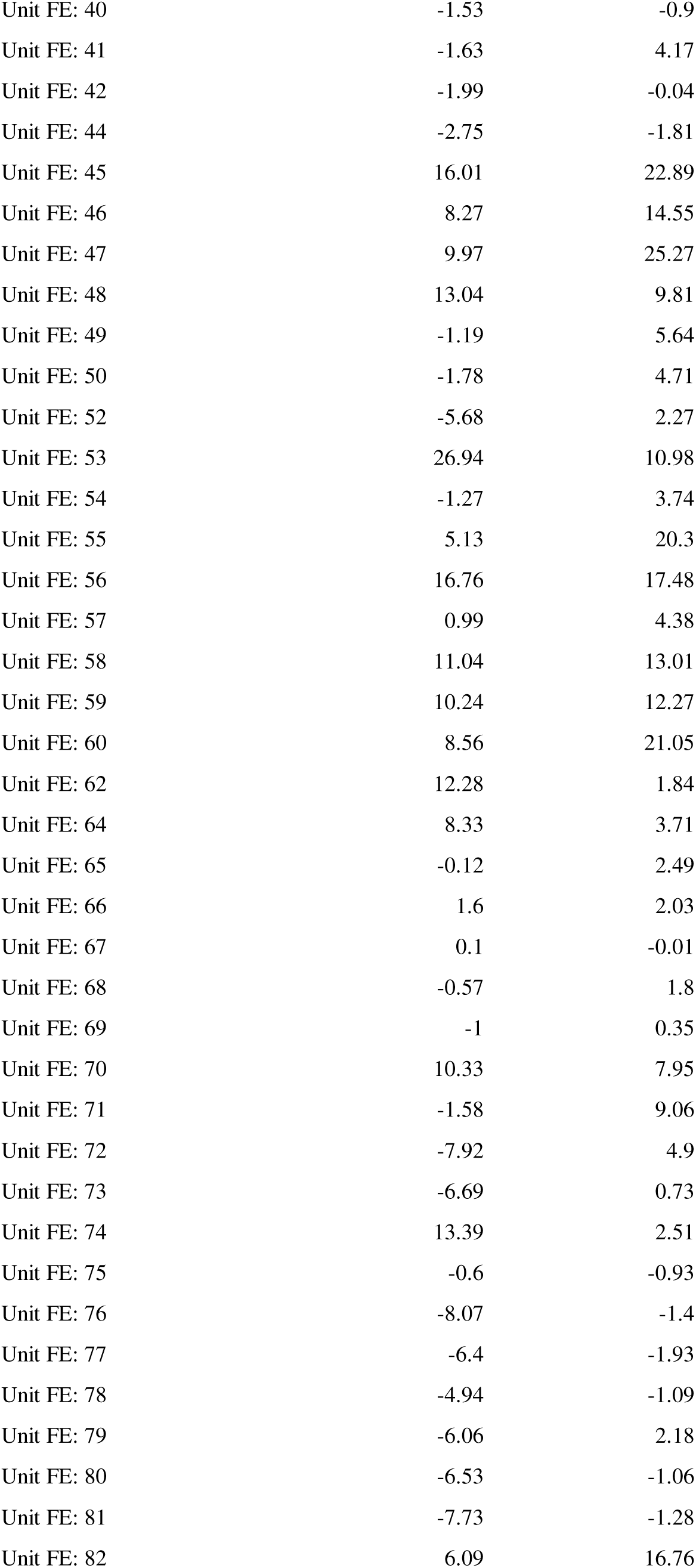
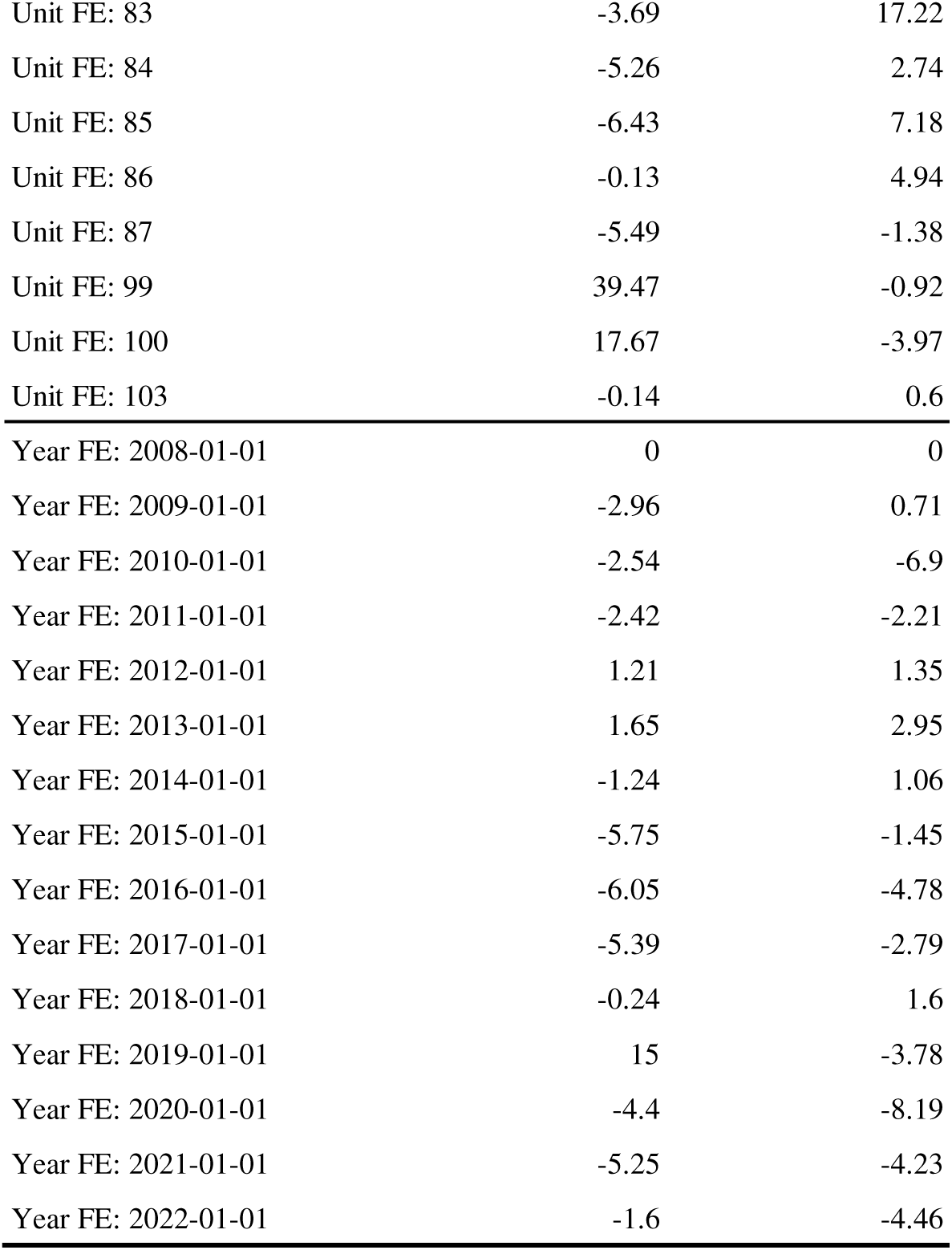
Dengue and leishmaniasis incidence long-differences estimation (Equation 2) fixed effects. Corresponds to the first and fourth columns in Table 1 in the main text. Note that these are analytically absorbed prior to estimation (using the Frisch–Waugh– Lovell theorem), so they have no standard errors.

| Term | Dengue Intercept | Leish Intercept |
| --- | --- | --- |
| Unit FE: 1 | 20.65 | -1.67 |
| Unit FE: 2 | 18.23 | -2.47 |
| Unit FE: 3 | -4.07 | -1.31 |
| Unit FE: 4 | 20.76 | 1.79 |
| Unit FE: 5 | -3.96 | -5.62 |
| Unit FE: 6 | -3.9 | -4.17 |
| Unit FE: 7 | -1.53 | -6.88 |
| Unit FE: 8 | -3.62 | -5.73 |
| Unit FE: 9 | 11.81 | -2.9 |
| Unit FE: 10 | -8.39 | -1.2 |
| Unit FE: 12 | -4.52 | -2.88 |
| Unit FE: 13 | 8.68 | -3.74 |
| Unit FE: 14 | 17.63 | -3.55 |
| Unit FE: 15 | 0.94 | -2.91 |
| Unit FE: 16 | -8.77 | -1.57 |
| Unit FE: 17 | -7.97 | 1.06 |
| Unit FE: 18 | -8.19 | -1.01 |
| Unit FE: 19 | 33.31 | -1.15 |
| Unit FE: 20 | -3.01 | -2.3 |
| Unit FE: 21 | -1.74 | -2.05 |
| Unit FE: 24 | -7.84 | 1.11 |
| Unit FE: 26 | -2.42 | -2.66 |
| Unit FE: 27 | -8.83 | 1.79 |
| Unit FE: 28 | -2.57 | 0.34 |
| Unit FE: 29 | -2.47 | -0.44 |
| Unit FE: 30 | -2.75 | -1.98 |
| Unit FE: 31 | -2.11 | -1.48 |
| Unit FE: 32 | -3.11 | 0.66 |
| Unit FE: 34 | 5.4 | 12.74 |
| Unit FE: 35 | -2.03 | -1.13 |
| Unit FE: 36 | -1.09 | -2.25 |
| Unit FE: 37 | 8.91 | 8.33 |
| Unit FE: 38 | 9.06 | 10.17 |
| Unit FE: 39 | -2.32 | -3.44 |
| Unit FE: 40 | -1.53 | -0.9 |
| Unit FE: 41 | -1.63 | 4.17 |
| Unit FE: 42 | -1.99 | -0.04 |
| Unit FE: 44 | -2.75 | -1.81 |
| Unit FE: 45 | 16.01 | 22.89 |
| Unit FE: 46 | 8.27 | 14.55 |
| Unit FE: 47 | 9.97 | 25.27 |
| Unit FE: 48 | 13.04 | 9.81 |
| Unit FE: 49 | -1.19 | 5.64 |
| Unit FE: 50 | -1.78 | 4.71 |
| Unit FE: 52 | -5.68 | 2.27 |
| Unit FE: 53 | 26.94 | 10.98 |
| Unit FE: 54 | -1.27 | 3.74 |
| Unit FE: 55 | 5.13 | 20.3 |
| Unit FE: 56 | 16.76 | 17.48 |
| Unit FE: 57 | 0.99 | 4.38 |
| Unit FE: 58 | 11.04 | 13.01 |
| Unit FE: 59 | 10.24 | 12.27 |
| Unit FE: 60 | 8.56 | 21.05 |
| Unit FE: 62 | 12.28 | 1.84 |
| Unit FE: 64 | 8.33 | 3.71 |
| Unit FE: 65 | -0.12 | 2.49 |
| Unit FE: 66 | 1.6 | 2.03 |
| Unit FE: 67 | 0.1 | -0.01 |
| Unit FE: 68 | -0.57 | 1.8 |
| Unit FE: 69 | -1 | 0.35 |
| Unit FE: 70 | 10.33 | 7.95 |
| Unit FE: 71 | -1.58 | 9.06 |
| Unit FE: 72 | -7.92 | 4.9 |
| Unit FE: 73 | -6.69 | 0.73 |
| Unit FE: 74 | 13.39 | 2.51 |
| Unit FE: 75 | -0.6 | -0.93 |
| Unit FE: 76 | -8.07 | -1.4 |
| Unit FE: 77 | -6.4 | -1.93 |
| Unit FE: 78 | -4.94 | -1.09 |
| Unit FE: 79 | -6.06 | 2.18 |
| Unit FE: 80 | -6.53 | -1.06 |
| Unit FE: 81 | -7.73 | -1.28 |
| Unit FE: 82 | 6.09 | 16.76 |
| Unit FE: 83 | -3.69 | 17.22 |
| Unit FE: 84 | -5.26 | 2.74 |
| Unit FE: 85 | -6.43 | 7.18 |
| Unit FE: 86 | -0.13 | 4.94 |
| Unit FE: 87 | -5.49 | -1.38 |
| Unit FE: 99 | 39.47 | -0.92 |
| Unit FE: 100 | 17.67 | -3.97 |
| Unit FE: 103 | -0.14 | 0.6 |
| Year FE: 2008-01-01 | 0 | 0 |
| Year FE: 2009-01-01 | -2.96 | 0.71 |
| Year FE: 2010-01-01 | -2.54 | -6.9 |
| Year FE: 2011-01-01 | -2.42 | -2.21 |
| Year FE: 2012-01-01 | 1.21 | 1.35 |
| Year FE: 2013-01-01 | 1.65 | 2.95 |
| Year FE: 2014-01-01 | -1.24 | 1.06 |
| Year FE: 2015-01-01 | -5.75 | -1.45 |
| Year FE: 2016-01-01 | -6.05 | -4.78 |
| Year FE: 2017-01-01 | -5.39 | -2.79 |
| Year FE: 2018-01-01 | -0.24 | 1.6 |
| Year FE: 2019-01-01 | 15 | -3.78 |
| Year FE: 2020-01-01 | -4.4 | -8.19 |
| Year FE: 2021-01-01 | -5.25 | -4.23 |
| Year FE: 2022-01-01 | -1.6 | -4.46 |

**Supplementary Table S6:** Dengue and leishmaniasis incidence long-differences estimation results (Equation 2) using *glmmTMB* (rather than *fixest*). Corresponds to Table 1 in main text. Main estimates match closely, but standard errors are larger. This is expected. In *fixest*, fixed effects are absorbed analytically using the Frisch–Waugh–Lovell theorem, which reduces dimensionality and isolates within-group variation. In *glmmTMB*, fixed effects are estimated explicitly, which introduces a large number of additional parameters (shown in **Supplementary Table S6** below). This increases model complexity and standard errors due to multicollinearity and estimating additional parameters. Yearly models include unit and year fixed effects, and biannual models include unit and biannual fixed effects. Standard errors are displayed in parentheses next to each estimated coefficient but are not clustered because *glmmTMB* does not offer clustering functionality. P-values indicating a statistical significance level below 0.05 are designated with an asterisk (*).

|  | Dengue<br>Yearly | Dengue<br>Biannual<br>Dry | Dengue<br>Biannual<br>Rainy | Leish Yearly | Leish<br>Biannual<br>Dry | Leish<br>Biannual<br>Rainy |
| --- | --- | --- | --- | --- | --- | --- |
| 5km x Post-2008 Yearly | 6.91* (3.35) |  |  | -0.38 (1.05) |  |  |
| 5km x Post-2008<br>Biannual |  | 2.06 (1.14) | 4.13* (2.07) |  | -0.20 (0.65) | 0.29 (0.65) |
| Urban | -1.10 (0.71) | -0.50* (0.24) | -0.72 (0.42) | 0.09 (0.22) | 0.04 (0.14) | 0.01 (0.13) |
| Agriculture | 0.01 (0.16) | 0.03 (0.05) | -0.03 (0.09) | -0.05 (0.05) | -0.05 (0.03) | -0.04 (0.03) |
| Precipitation | 5.49 (12.00) | 11.78 (9.61) | 6.70 (8.46) | -10.10* (3.74) | -15.46* (5.49) | -2.81 (2.66) |
| Temperature | 6.80 (7.70) | 1.53 (1.68) | 4.32 (4.74) | 7.64* (2.40) | 2.30* (0.96) | 1.78 (1.49) |
| Temperature^2 | 0.45 (1.06) | 0.04 (0.30) | 0.06 (0.62) | -0.06 (0.33) | -0.23 (0.17) | 0.00 (0.20) |
| FEs |  |  |  |  |  |  |
| Unit | X | X | X | X | X | X |
| Year | X |  |  | X |  |  |
| Biannual |  | X | X |  | X | X |

**Supplementary Table S7:**
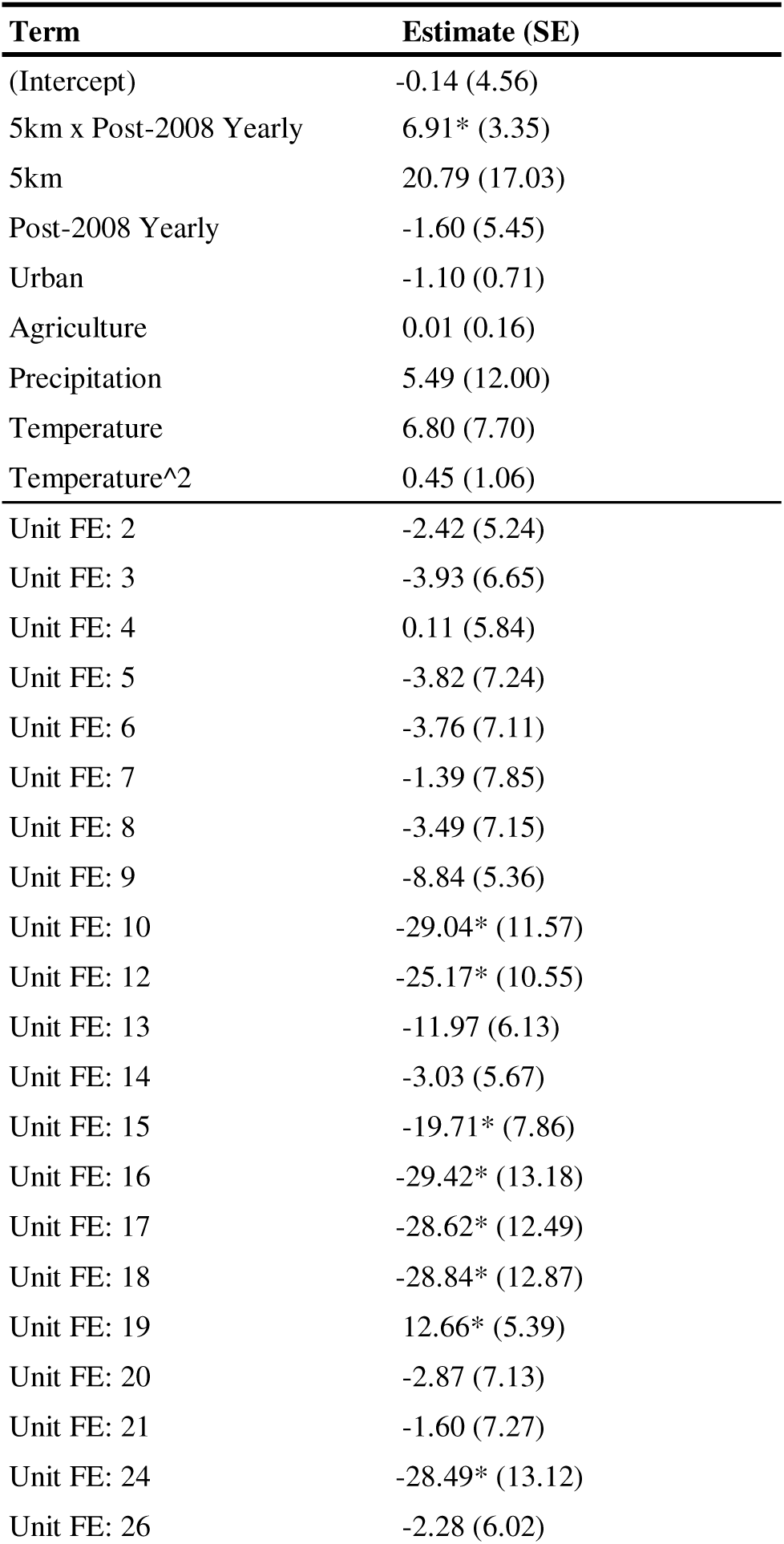
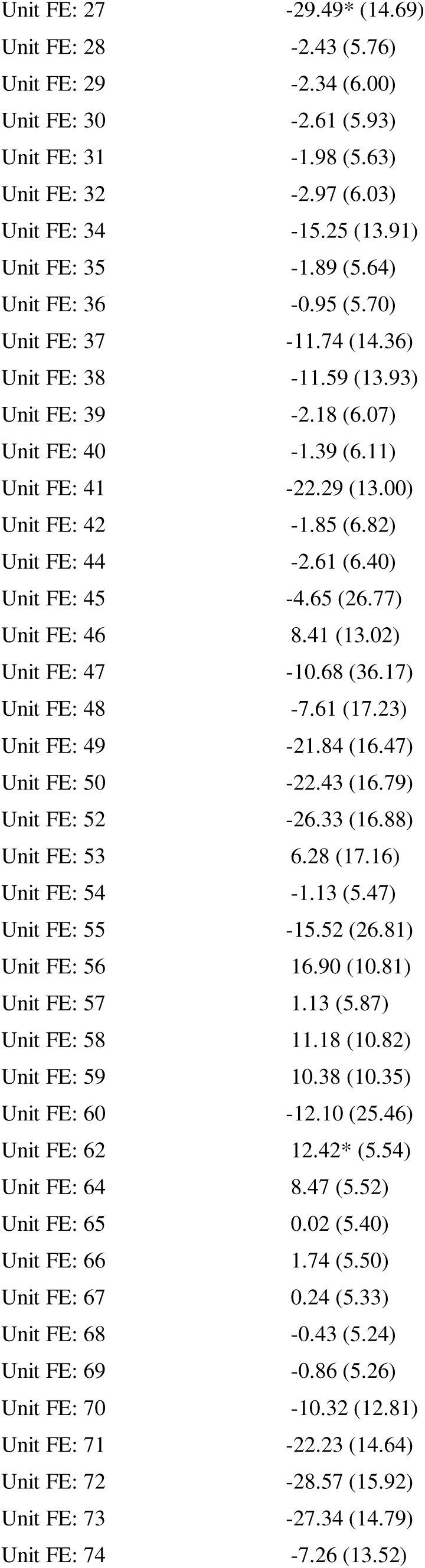
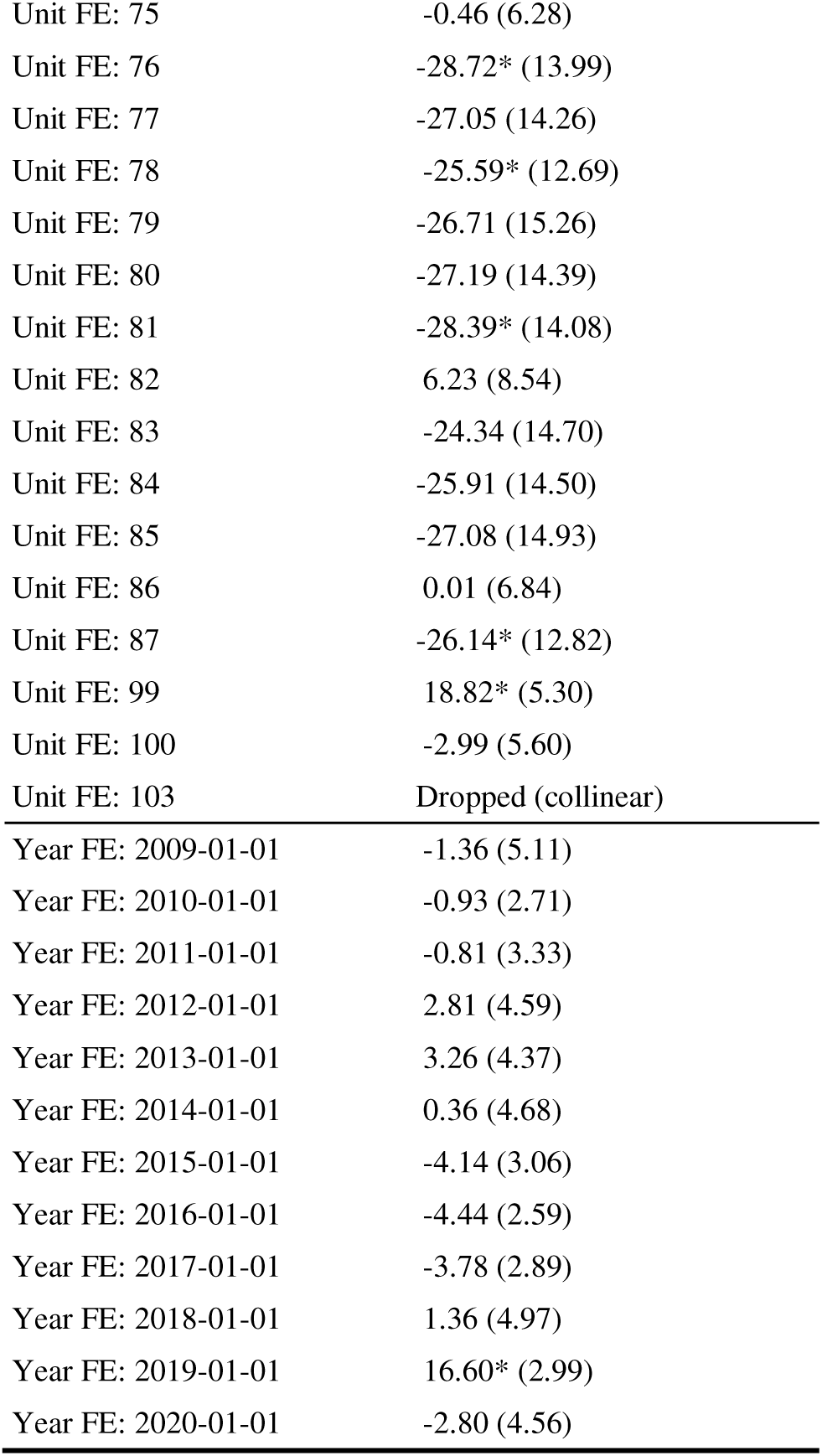
Dengue incidence long-differences estimation results (Equation 2) using *glmmTMB* (rather than *fixest*) with all interaction and intercept terms included. Corresponds to the first column of **Supplementary Table S6**. Model includes unit and year fixed effects. Standard errors are displayed in parentheses next to each estimated coefficient but are not clustered because *glmmTMB* does not offer clustering functionality. P-values indicating a statistical significance level below 0.05 are designated with an asterisk (*). While these values are somewhat conceptually similar to those reported in **Supplementary Table S4**, they are estimated differently. Fixed effect intercepts from *fixest* represent group-specific means calculated without shrinkage. In contrast, random effects from *glmmTMB* are estimated as deviations from the global mean and are partially pooled and shrunk toward zero.

## Supplementary Figures S1–S10

**Supplementary Figure S1:**
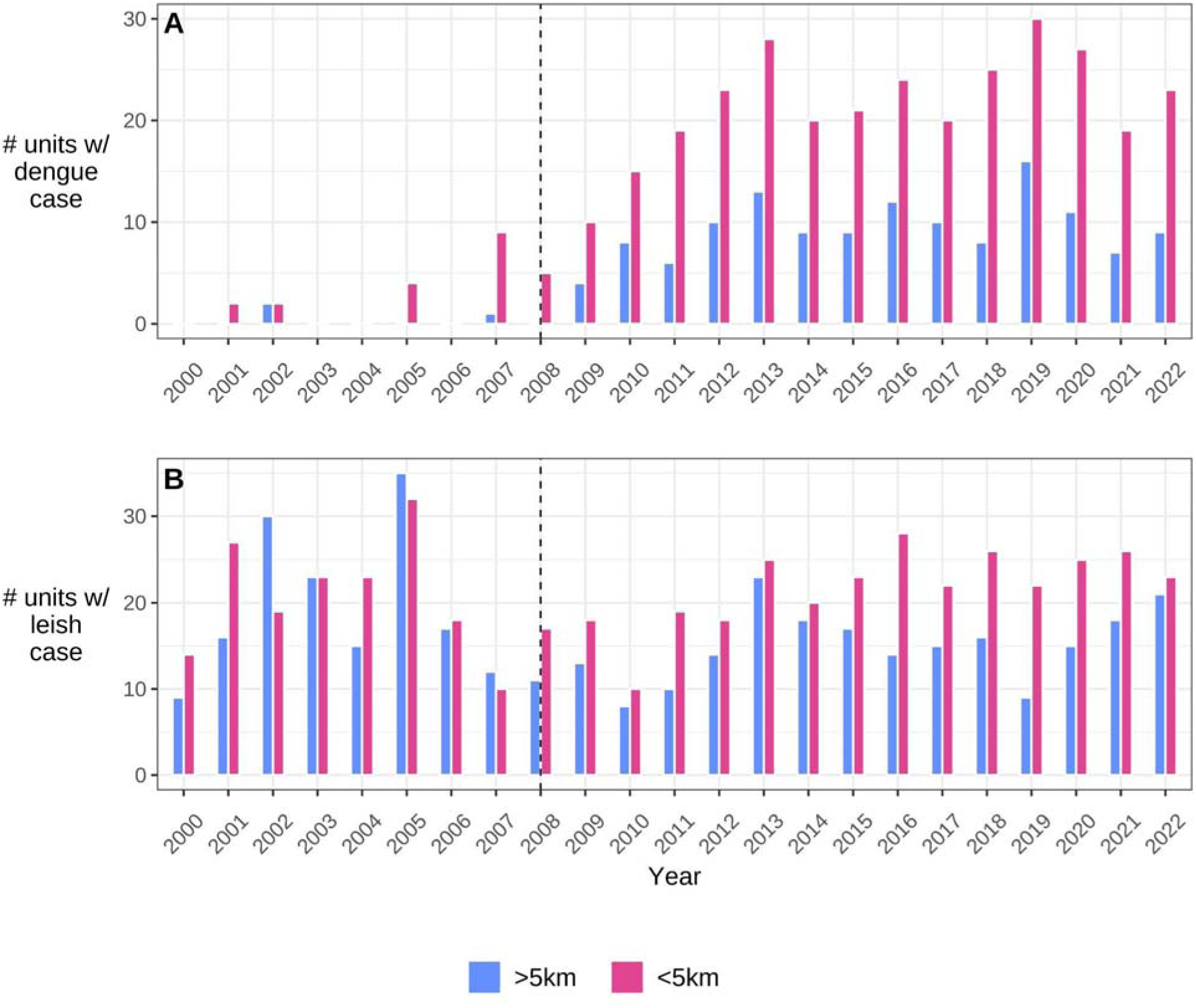
Bar plots displaying disease case data coverage: number of healthcare facilities (i.e., spatial units) reporting at least one dengue (A) or leishmaniasis (B) case by year and exposure group (color, where blue units are considered unexposed and pink units exposed). Buffer facilities are included in this plot to display dengue dynamics throughout the region’s main transportation network (i.e., facilities >5km and <10km from the highway are displayed here but not included in the main model specification). Facilities disconnected from the main transit network remain excluded (grey points in **Figure 1**). Prior to paving, dengue was reported in healthcare facilities in Puerto Maldonado, Iberia, Mavila, Mazuko, Iñapari, Planchon, Loero, Laberinto, and Boca Colorado.

**Supplementary Figure S2:**
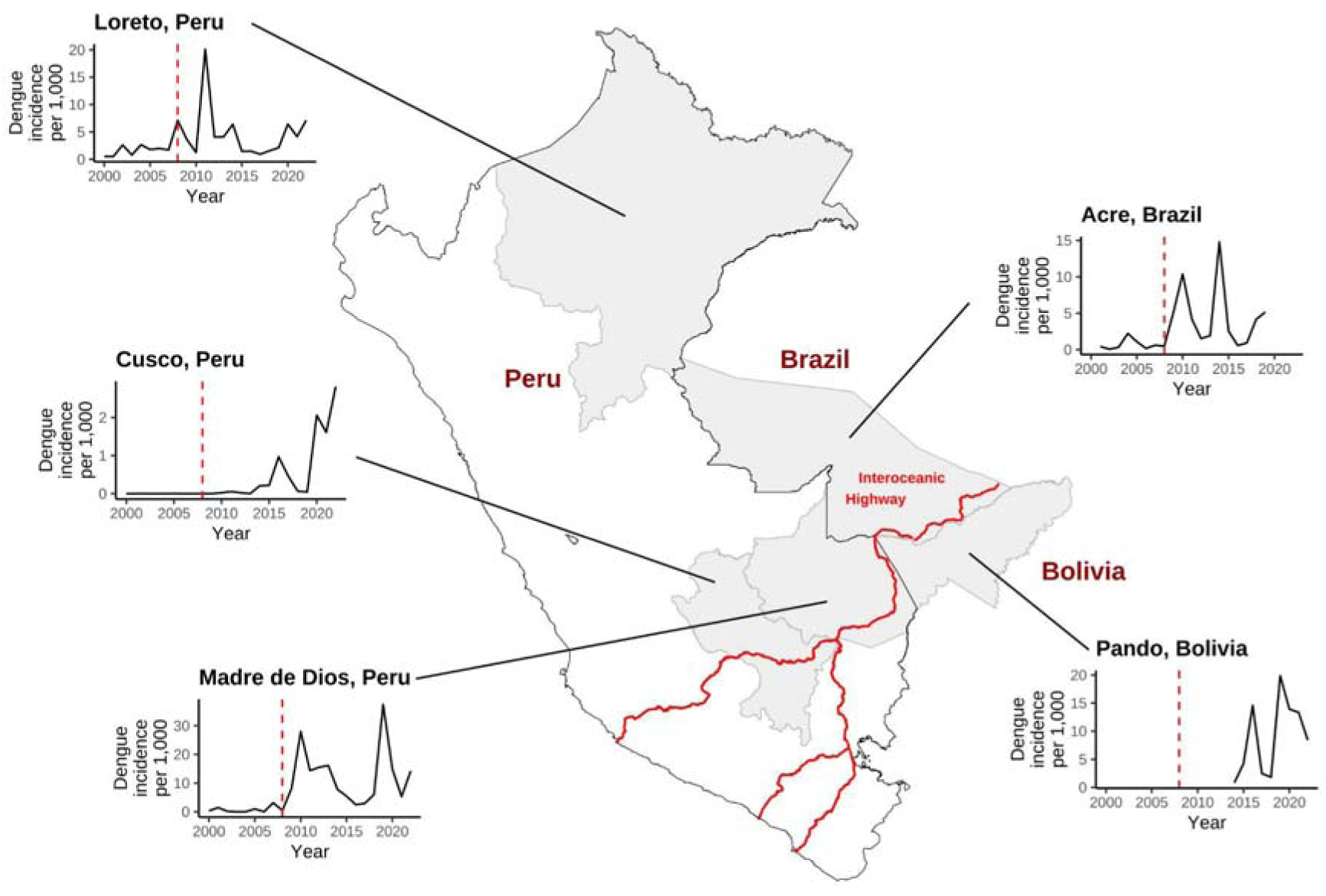
Time series plots of yearly dengue incidence rates per 1,000 for regions bordering and otherwise relevant to Madre de Dios^10–12^. Interoceanic Highway colored in red. Clockwise from top right: Acre, Brazil, dengue-endemic pre-2009 and bordering to the Northeast; Pando, Bolivia, dengue-endemic pre-2009 and bordering to the East (no data are available pre-2014); Madre de Dios, Peru, study setting; Cusco, Peru, only a small portion of the department is suitable for dengue, bordering to the West; Loreto, Peru, dengue-endemic and the only other substantially populated Amazonian department in Peru, not bordering Madre de Dios.

**Supplementary Figure S3:**
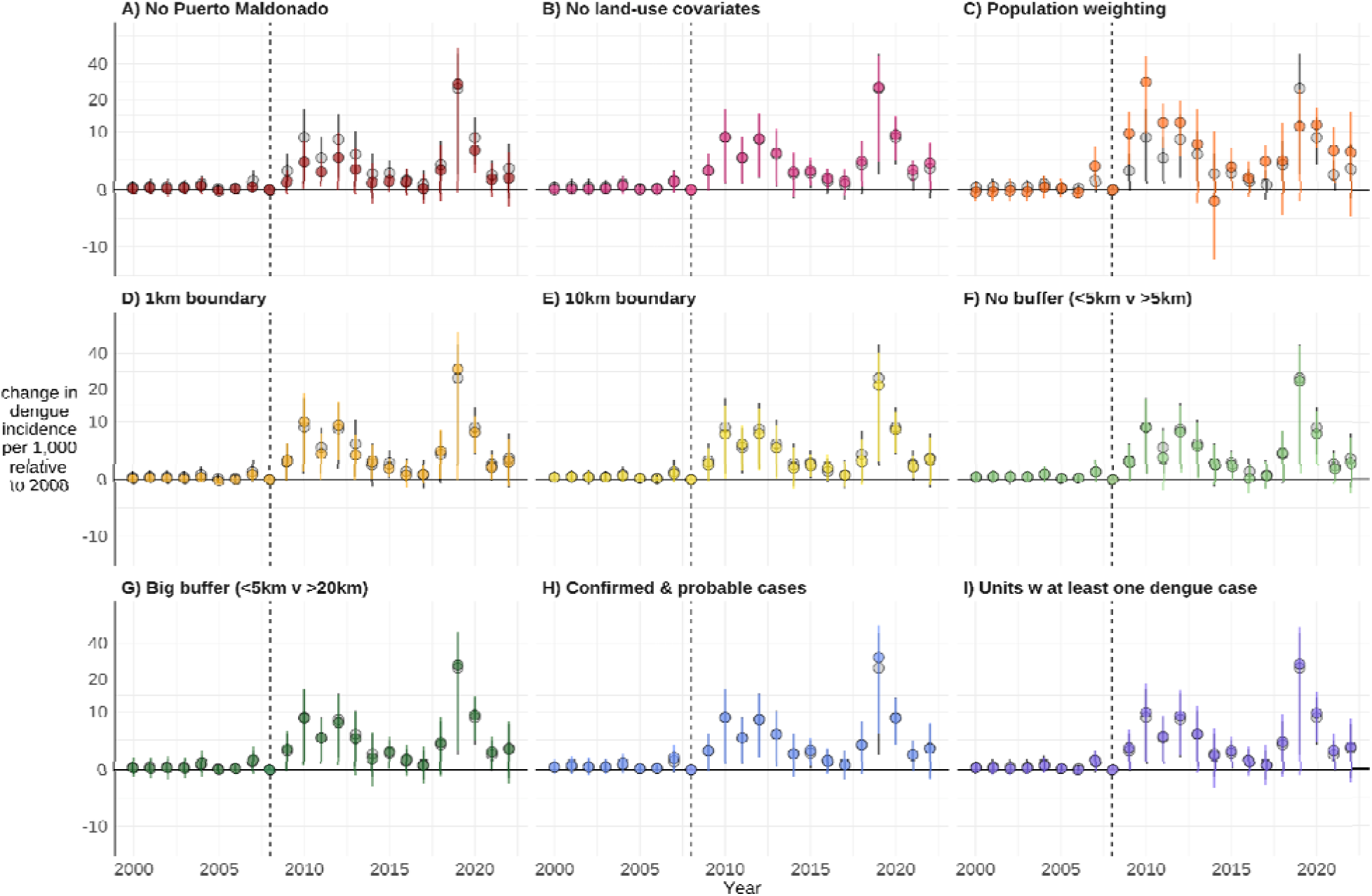
Model results are robust to various model specifications. Estimation results from Equation 1 for the following model specifications compared to the main model (shown in grey): A) model excluding Puerto Maldonado (the region’s largest city), B) model excluding land-use covariates, C) population-weighted model, D) model designating <1KM from the highway as exposed and >1km from the highway as unexposed, E) model designating <10KM as exposed and >10km as unexposed, F) model with no spatial buffer (<5km exposed, >5km unexposed), G) model with a larger spatial buffer (<5km exposed, >20km unexposed), H) model including both confirmed and probable cases, and I) model including only spatial units reporting at least one dengue case during the study period. Average treatment effect (points) with 95% confidence interval (error bars). Standard errors clustered by spatial group. Vertical dotted line at 2008 represents the last year before the highway was paved (baseline year). Note that the y-axis uses a log scale.

**Supplementary Figure S4:**
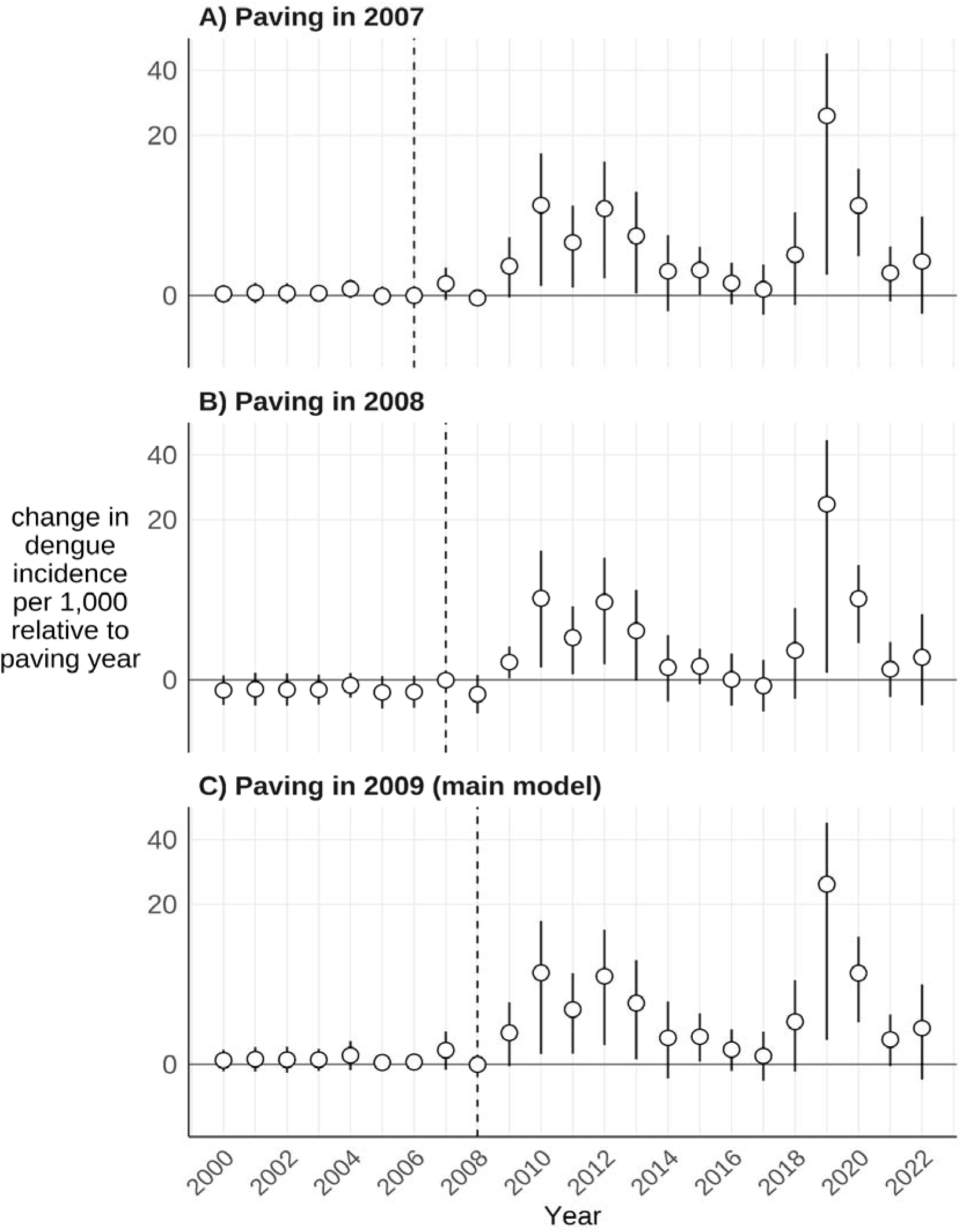
Model results are robust to exposure year designation. Estimation results from Equation 1 for exposure year designated as A) 2007, B) 2008, and C) 2009 (main model). Vertical dotted lines denote the baseline year, i.e., the last year before exposure: A) 2006, B) 2007, and C) 2008 (main model). Note that the y-axis uses a log scale.

**Supplementary Figure S5:**
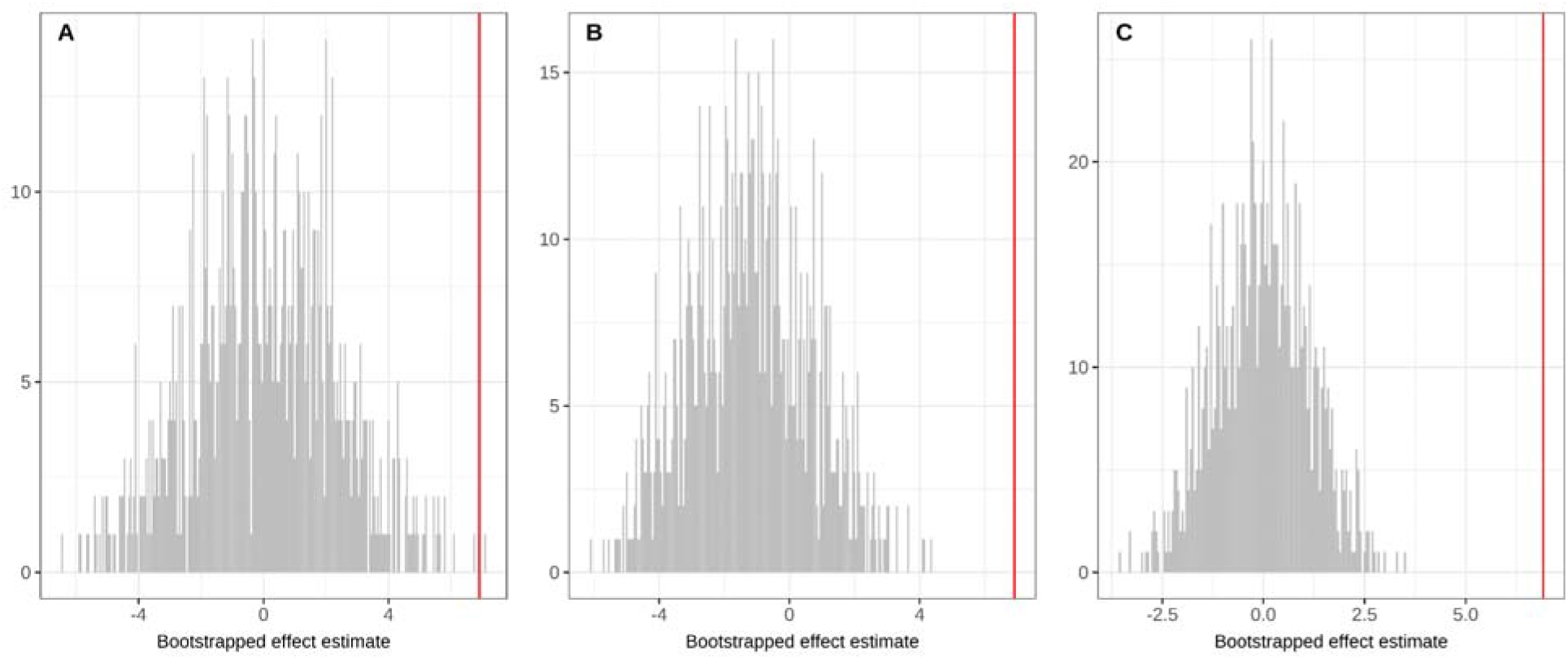
Permutation inference analysis. Histograms of bootstrapped effect estimates from three methods of randomly shuffling the exposure variable (grey bars) compared to the estimated effect from the non-bootstrapped sample (vertical red lines on the right of the panels). The three methods are: A) Random, the entire exposure column is randomly shuffled across facilities and years, B) Full, randomly assign facilities to exposure, maintaining temporal structure, and C) Within, randomly assign years as treated within facilities, maintaining spatial structure. In all cases, histograms are centered on zero effect and the distributions are well below the empirically estimated effect from the main model, indicating that the effect is unlikely to be confounded by chance spatial or temporal structure in the data.

**Supplementary Figure S6:**
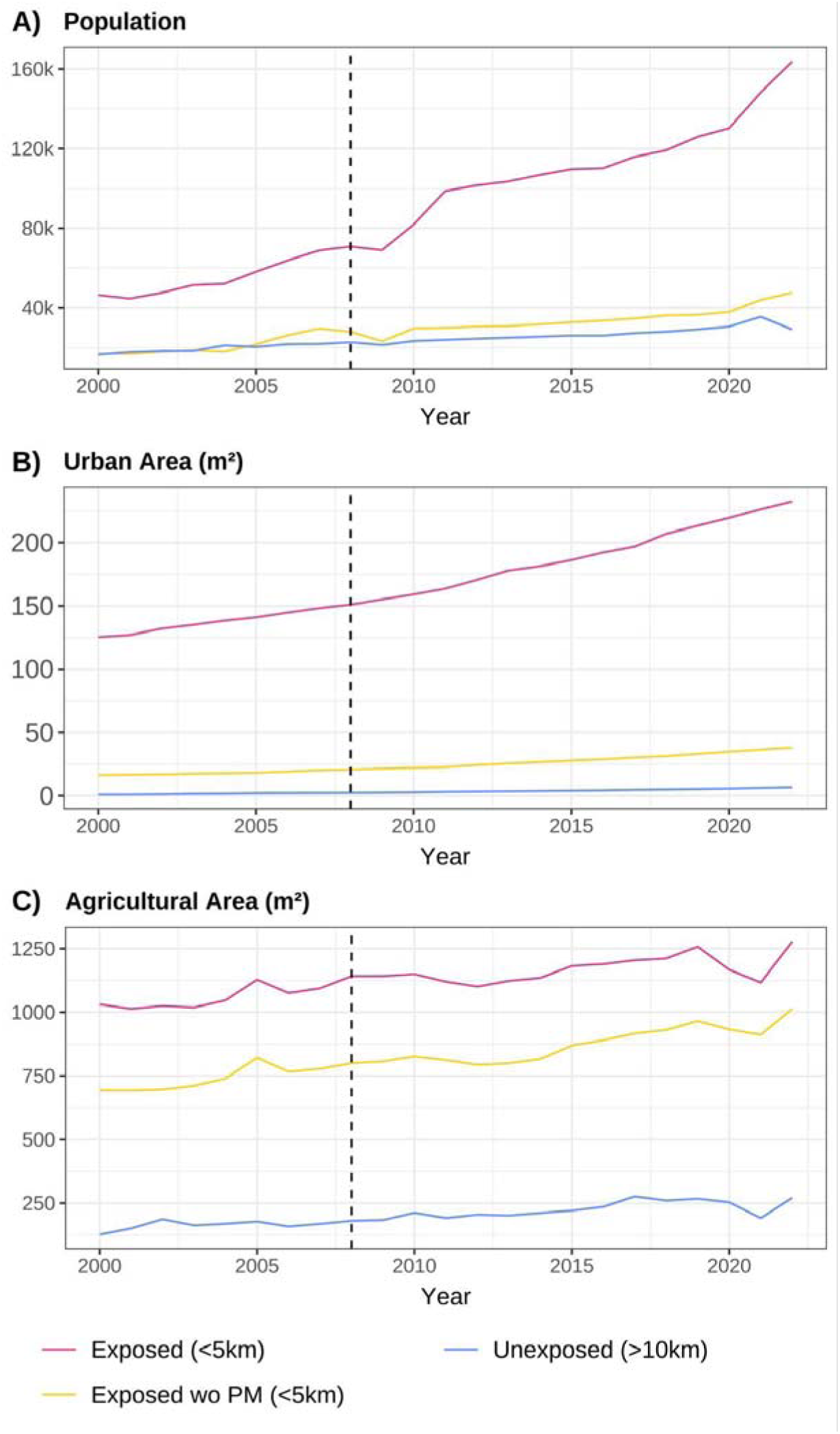
Time series plots displaying yearly measures of population, urban area, and agricultural area by exposure group (color). Lines display the ‘exposed’ group (pink), the exposed group excluding Puerto Maldonado (PM, yellow), the largest city in the region, and the unexposed group (blue). Sources and calculations of these plots are discussed in the Methods section and in the Supplementary Information.

**Supplementary Figure S7:**
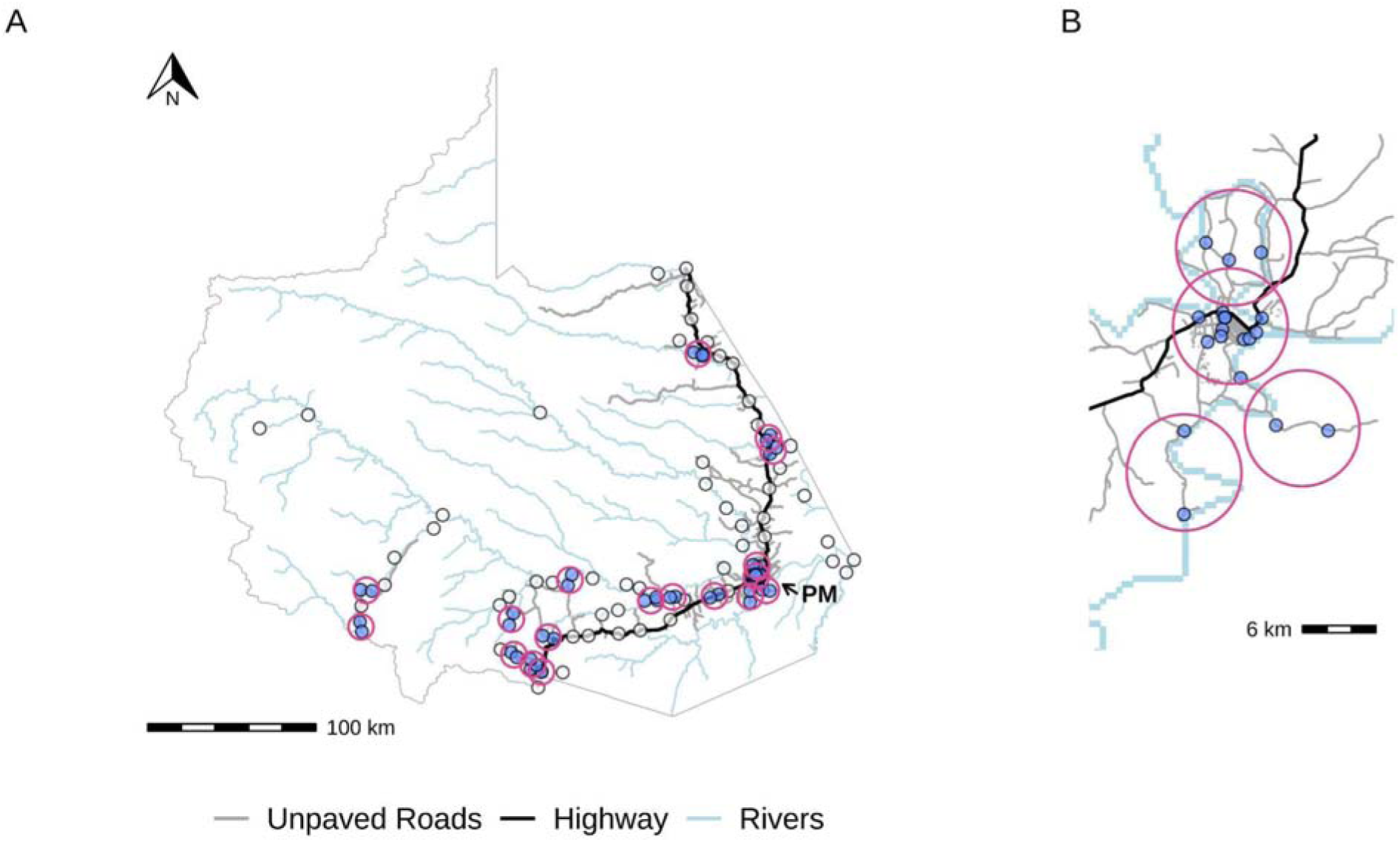
All healthcare facilities (n = 102) mapped onto the study region (A), with a zoomed view of Puerto Maldonado shown in the inset (B). Points are colored blue if they are spatially grouped and their standard errors are clustered (discussed in the **Methods** section). Points are colored white if they have sufficient spatial separation and are designated as their own group. Pink circles designate groupings.

**Supplementary Figure S8:**
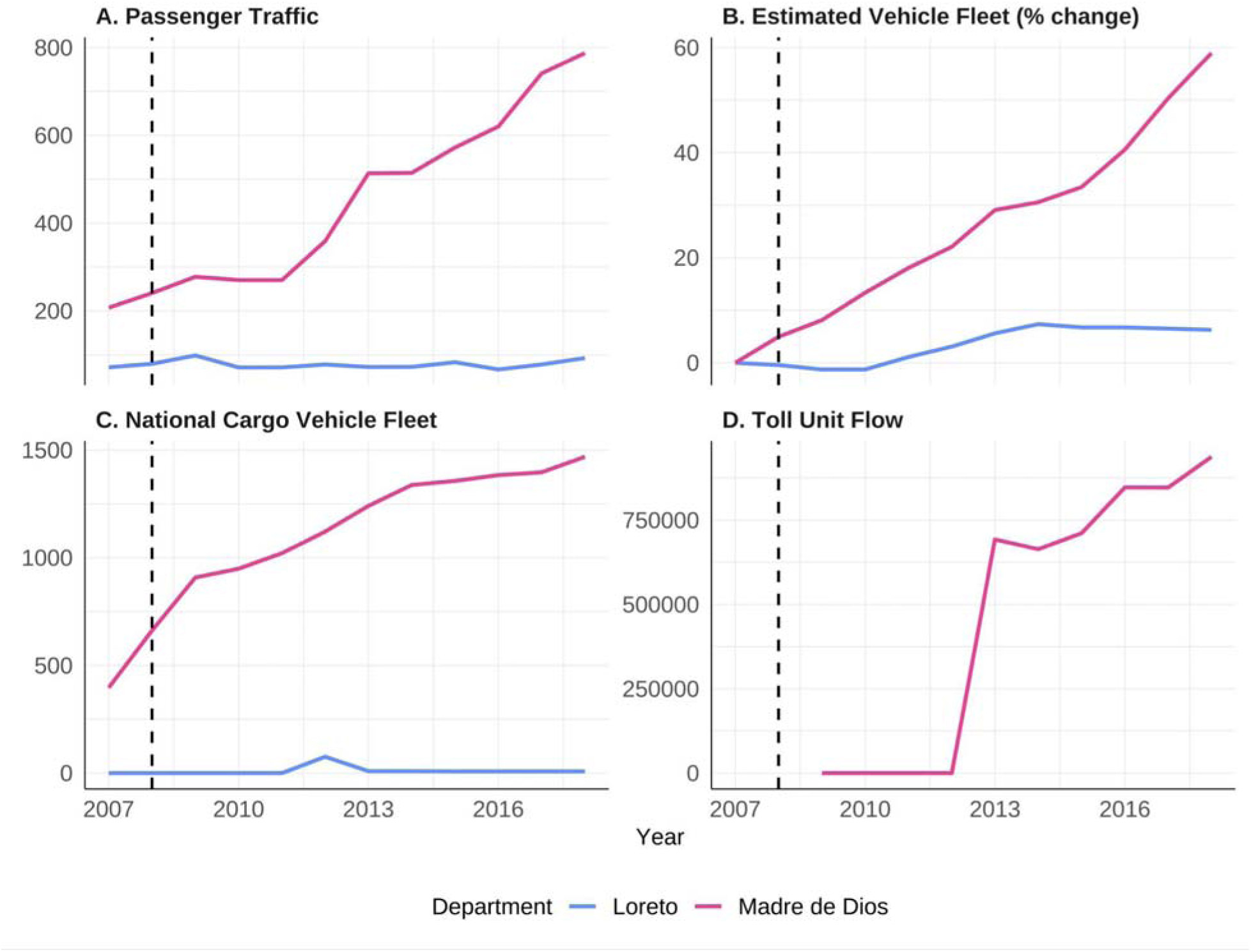
Supplementary traffic volume data shows increases across multiple metrics post highway paving in Madre de Dios, Peru. Official Peruvian governmental traffic volumes (passenger traffic [A], estimated general vehicle fleet [B], national cargo vehicle fleet [C], and toll unit flow [D])^13^ are reported for both Madre de Dios, Peru (pink, study region) and Loreto, Peru (blue, the only other substantially populated Amazonian department in Peru, which did not undergo highway construction during the study period, see map in **Supplementary Figure S2**). Notes on Panel D: toll unit flow was not available for Loreto throughout the study period, and toll unit flow data was not available for Madre de Dios until 2009 and reported as zero until 2013. Vertical dotted line at 2008 representing the last year before the highway was paved (baseline year).

**Supplementary Figure S9:**
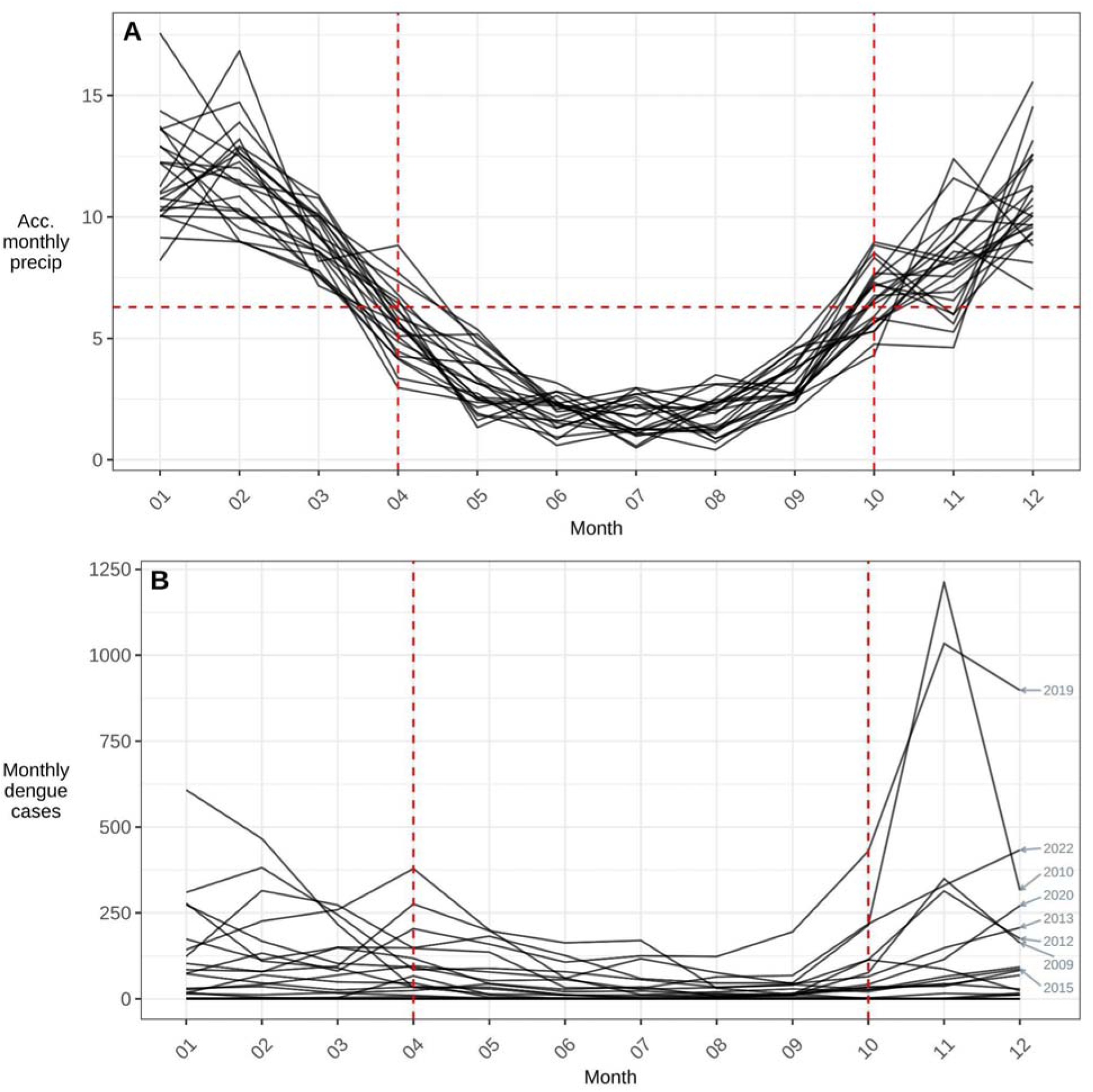
Seasonality of monthly accumulated precipitation (A) and dengue cases (B) in Madre de Dios, Peru during the study period (2000–2022). The red horizontal line in panel A shows the average monthly accumulated precipitation across the entirety of the study period. The red vertical lines show the months that intersect with the red horizontal line delineating rainy (October – March) versus dry season (April – September).

**Supplementary Figure S10:**
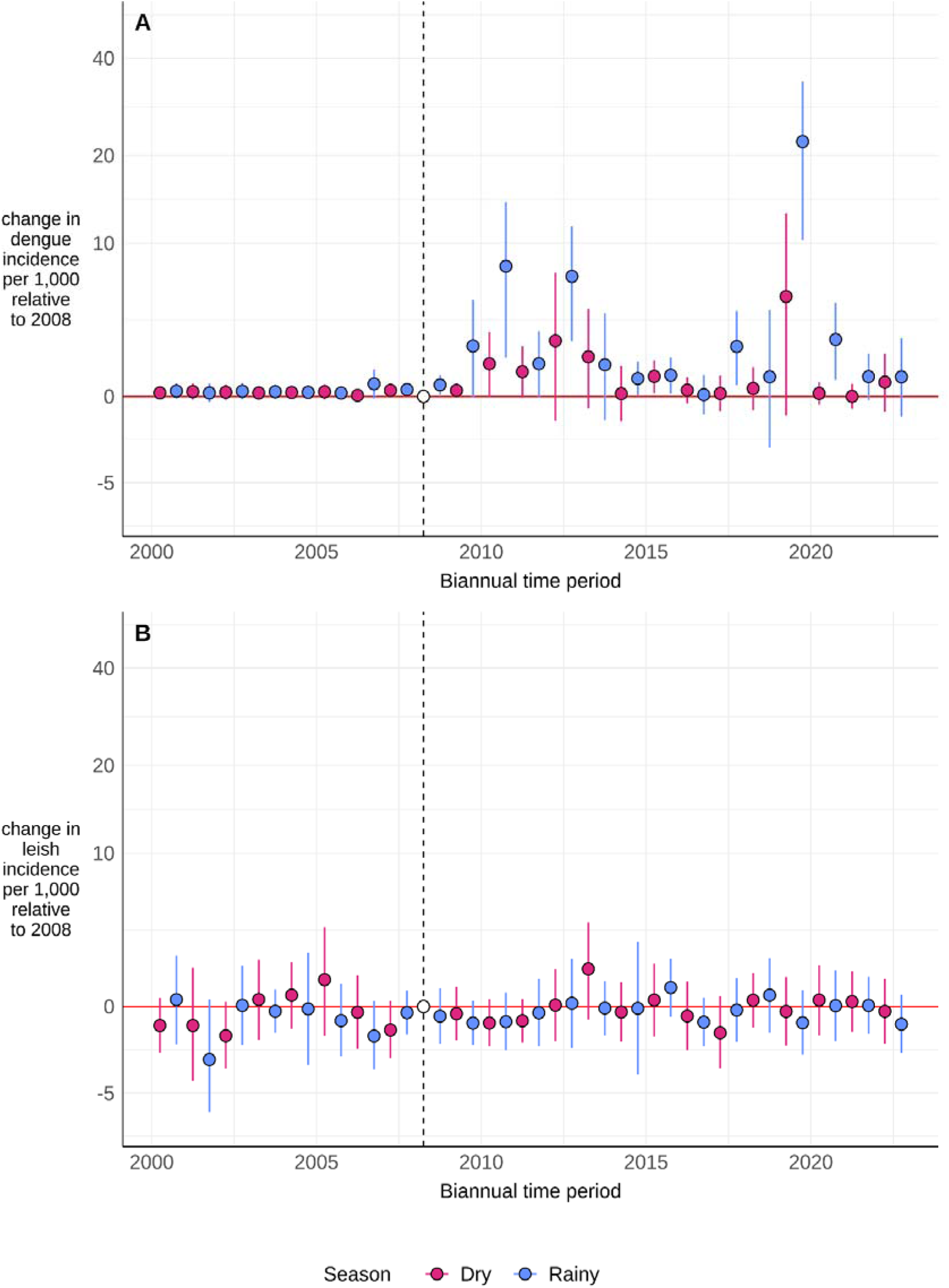
Disaggregated biannual model results for dengue (A) and leishmaniasis (B) from Equation 1, colored by season (rainy [red] vs. dry [blue]). Yearly treatment effects are represented by points, and 95% confidence intervals are represented by error bars (standard errors clustered by spatial group). The vertical dotted line at 2008 represents the last year before the highway was paved (baseline year). Note that the y-axis uses a log scale.

## References

1. Thacker, S. et al. Infrastructure for sustainable development. Nat. Sustain. 2, 324–331 (2019).

2. Morgan, R. K. Environmental impact assessment: the state of the art. Impact Assess. Proj. Apprais. 30, 5–14 (2012).

3. Patz, J. A. et al. Unhealthy Landscapes: Policy Recommendations on Land Use Change and Infectious Disease Emergence. Environ. Health Perspect. 112, 1092–1098 (2004).

4. Vilela, T. et al. A better Amazon road network for people and the environment. Proc. Natl. Acad. Sci. 117, 7095–7102 (2020).

5. Lana, R. M., Gomes, M. F. da C., Lima, T. F. M. de, Honório, N. A. & Codeço, C. T. The introduction of dengue follows transportation infrastructure changes in the state of Acre, Brazil: A network-based analysis. PLoS Negl. Trop. Dis. 11, e0006070 (2017).

6. Li, Q., Cao, W., Ren, H., Ji, Z. & Jiang, H. Spatiotemporal responses of dengue fever transmission to the road network in an urban area. Acta Trop. 183, 8–13 (2018).

7. Buckee, C., Noor, A. & Sattenspiel, L. Thinking clearly about social aspects of infectious disease transmission. Nature 595, 205–213 (2021).

8. Wesolowski, A. et al. Multinational patterns of seasonal asymmetry in human movement influence infectious disease dynamics. Nat. Commun. 8, 2069 (2017).

9. Scullion, J. J., Vogt, K. A., Sienkiewicz, A., Gmur, S. J. & Trujillo, C. Assessing the influence of land-cover change and conflicting land-use authorizations on ecosystem conversion on the forest frontier of Madre de Dios, Peru. Biol. Conserv. 171, 247–258 (2014).

10. Salmón-Mulanovich, G. et al. Individual and Spatial Risk of Dengue Virus Infection in Puerto Maldonado, Peru. Am. J. Trop. Med. Hyg. 99, 1440–1450 (2018).

11. Jensen, K. E., et al. Small scale migration along the interoceanic highway in Madre de Dios, Peru: an exploration of community perceptions and dynamics due to migration. BMC Int. Health Hum. Rights 18, 12 (2018).

12. Sánchez-Cuervo, A. M. et al. Twenty years of land cover change in the southeastern Peruvian Amazon: implications for biodiversity conservation. Reg. Environ. Change 20, 8 (2020).

13. Oliveira, A. S. et al. Bringing economic development for whom? An exploratory study of the impact of the Interoceanic Highway on the livelihood of smallholders in the Amazon. Landsc. Urban Plan. 188, 171–179 (2019).

14. Riley-Powell, A. R. et al. The Impact of Road Construction on Subjective Well-Being in Communities in Madre de Dios, Peru. Int. J. Environ. Res. Public. Health 15, 1271 (2018).

15. Sanchez, J. F. et al. Unstable Malaria Transmission in the Southern Peruvian Amazon and Its Association with Gold Mining, Madre de Dios, 2001–2012. Am. J. Trop. Med. Hyg. 96, 304–311 (2017).

16. Monath, T. P. Dengue: the risk to developed and developing countries. Proc. Natl. Acad. Sci. 91, 2395–2400 (1994).

17. WHO. Dengue: the fastest growing mosquito-borne disease in the world. https://www.who.int/news/item/29-10-2010-dengue-the-fastest-growing-mosquito-borne-disease-in-the-world (2010).

18. Dengue - Global situation. https://www.who.int/emergencies/disease-outbreak-news/item/2024-DON518 (2024).

19. Guagliardo, S. A. et al. Patterns of Geographic Expansion of Aedes aegypti in the Peruvian Amazon. PLoS Negl. Trop. Dis. 8, e3033 (2014).

20. Stoddard, S. T., et al. The Role of Human Movement in the Transmission of Vector-Borne Pathogens. PLoS Negl. Trop. Dis. 3, e481 (2009).

21. Harrington, L. C. et al. Dispersal of the dengue vector Aedes aegypti within and between rural communities. Am. J. Trop. Med. Hyg. 72, 209–220 (2005).

22. Guthmann, J. P. et al. Patients’ associations and the control of leishmaniasis in Peru. Bull. World Health Organ. 75, 39–44 (1997).

23. Zorrilla, V., et al. Distribution and identification of sand flies naturally infected with Leishmania from the Southeastern Peruvian Amazon. PLoS Negl. Trop. Dis. 11, e0006029 (2017).

24. Morrison, A. C., Ferro, C., Morales, A., Tesh, R. B. & Wilson, M. L. Dispersal of the sand fly Lutzomyia longipalpis (Diptera: Psychodidae) at an endemic focus of visceral leishmaniasis in Colombia. J. Med. Entomol. 30, 427–435 (1993).

25. Saliba, E. K. & Oumeish, O. Y. Reservoir hosts of cutaneous leishmaniasis. Clin. Dermatol. 17, 275–277 (1999).

26. Soares, L., Abad-Franch, F. & Ferraz, G. Epidemiology of cutaneous leishmaniasis in central Amazonia: a comparison of sex-biased incidence among rural settlers and field biologists. Trop. Med. Int. Health 19, 988–995 (2014).

27. Dimick, J. B. & Ryan, A. M. Methods for Evaluating Changes in Health Care Policy: The Difference-in-Differences Approach. JAMA 312, 2401–2402 (2014).

28. Orba, Y. et al. Diverse mosquito-specific flaviviruses in the Bolivian Amazon basin. J. Gen. Virol. 102, 001518 (2021).

29. Moore, P. A decade on, Peru’s corruption-laden Interoceanic Highway troubles communities. Dialogo Chino https://dialogochino.net/en/infrastructure/52497-peru-corruption-interoceanic-highway-trouble-communities/ (2022).

30. Ministerio de Transportes y Comunicaciones. https://portal.mtc.gob.pe/estadisticas/transportes.html.

31. Liang, S. et al. Construction sites as an important driver of dengue transmission: implications for disease control. BMC Infect. Dis. 18, 382 (2018).

32. Dutta, P. et al. Distribution of potential dengue vectors in major townships along the national highways and trunk roads of northeast India. Southeast Asian J. Trop. Med. Public Health 29, 173–176 (1998).

33. Guagliardo, S. A. J., et al. The genetic structure of Aedes aegypti populations is driven by boat traffic in the Peruvian Amazon. PLoS Negl. Trop. Dis. 13, e0007552 (2019).

34. Changing dynamics of Aedes aegypti invasion and vector-borne disease risk for rural communities in the Peruvian Amazon | bioRxiv. https://www.biorxiv.org/content/10.1101/2024.09.04.611168v1.full.

35. Flynn, L., Bery, R. & Kaitano, A. E. Emerging Infectious Diseases and Impact Assessments. (2013).

36. Peru ends era of ‘roadless wilderness’ in its Amazon rainforests. The Independent https://www.independent.co.uk/climate-change/news/peru-amazon-rainforests-road-building-roadless-wilderness-madre-de-dios-a8196971.html (2018).

37. Kleinschroth, F. & Healey, J. R. Impacts of logging roads on tropical forests. Biotropica 49, 620–635 (2017).

38. Durand, S. et al. Cost of controlling the dengue vector Aedes aegypti in the Peruvian amazon. Rev. Peru. Med. Exp. Salud Pública 41, 46–53 (2024).

39. Dengue is raging in Brazil. A promising local vaccine is at least a year away. https://www.science.org/content/article/dengue-raging-brazil-promising-local-vaccine-least-year-away.

40. Lenharo, M. Brazil’s record dengue surge: why a vaccine campaign is unlikely to stop it. Nature 627, 250–251 (2024).

41. Morrison, A. C., et al. Potential for community based surveillance of febrile diseases: Feasibility of self-administered rapid diagnostic tests in Iquitos, Peru and Phnom Penh, Cambodia. PLoS Negl. Trop. Dis. 15, e0009307 (2021).

42. Valdivia-Conroy, B. et al. Diagnostic performance of the rapid test for the detection of NS1 antigen and IgM and IgG anti-antibodies against dengue virus. Rev. Peru. Med. Exp. Salud Pública 39, 434–441 (2023).

43. Vittor, A. Y. et al. The effect of deforestation on the human-biting rate of Anopheles darlingi, the primary vector of Falciparum malaria in the Peruvian Amazon. Am. J. Trop. Med. Hyg. 74, 3–11 (2006).

44. Eisenberg, J. N. S. et al. Environmental change and infectious disease: How new roads affect the transmission of diarrheal pathogens in rural Ecuador. Proc. Natl. Acad. Sci. 103, 19460–19465 (2006).

45. Gonzalez, A. et al. Human movement and transmission dynamics early in Ebola outbreaks. 2023.12.18.23300175 Preprint at 10.1101/2023.12.18.23300175 (2023).

46. Dirección Regional de Salud Madre de Dios - Diresa Madre de Dios. https://www.gob.pe/regionmadrededios-diresa (2021).

47. Castagnetto, J. ubigeo. https://github.com/jmcastagnetto/ubigeo/?tab=readme-ov-file (2021).

48. Plataforma Nacional de Datos Abiertos. Plataforma Nacional de Datos Abiertos https://www.datosabiertos.gob.pe/dataset/establecimientos-de-salud (2024).

49. WorldPop. WorldPop :: Population Counts. https://hub.worldpop.org/project/categories?id=3 (2023).

50. Mapbiomas Brasil. https://mapbiomas.org/.

51. Muñoz Sabater, J. ERA5-Land monthly averaged data from 1950 to present. Copernicus Climate Change Service (C3S) Climate Data Store (CDS) https://cds.climate.copernicus.eu/cdsapp#!/dataset/10.24381/cds.68d2bb30?tab=overview (2019).

52. Peru’s Interoceanic: the Most Corrupt Highway in the World. Upper Amazon Conservancy (EN) https://www.upperamazon.org/news/fz8vmhz9ytujotmqksrxoex5rljfcv-mnsk7 (2018).

53. UNASUR - COSIPLAN. https://www.iirsa.org/proyectos/detalle_proyecto.aspx?h=319 (2017).

54. Bravo Orellana, S. Carretera Interoceánica Sur del Perú. Retos e innovación. (2013).

55. Oficina de Estadística - OGPP & Ministerio de Transportes y Comunicaciones. MAPA VIAL DEL PERU. (2007).

56. Oficina de Estadística & Oficina General de Planeamiento y Presupuesto - MTC. MAPA VIAL - MADRE DE DIOS. (2009).

57. Johanson, M. A highway in the Amazon ushers in a new era of deforestation and social upheaval. World Wildlife Fund https://www.worldwildlife.org/stories/how-the-interoceanic-highway-ushered-in-a-new-era-of-deforestation-and-social-upheaval-in-the-amazon (2024).

58. Perz, S. G., Mendoza, E. R. H. & Dos Santos Pimentel, A. Seeing the broader picture: Stakeholder contributions to understanding infrastructure impacts of the Interoceanic Highway in the southwestern Amazon. World Dev. 159, 106061 (2022).

59. Carrasco-Escobar, G., Manrique, E., Tello-Lizarraga, K. & Miranda, J. J. Travel Time to Health Facilities as a Marker of Geographical Accessibility Across Heterogeneous Land Coverage in Peru. Front. Public Health 8, (2020).

60. Mordecai, E. A. et al. Thermal biology of mosquito-borne disease. Ecol. Lett. 22, 1690–1708 (2019).

61. Berge, L., Krantz, S., McDermott, G. & Lenth, R. fixest: Fast Fixed-Effects Estimations. (2024).

62. Chen, J. & Roth, J. Logs with Zeros? Some Problems and Solutions. Q. J. Econ. 139, 891–936 (2024).

63. Brooks, M. et al. glmmTMB: Generalized Linear Mixed Models using Template Model Builder. (2025).

64. Rockhill, B., Newman, B. & Weinberg, C. Use and misuse of population attributable fractions. Am. J. Public Health 88, 15–19 (1998).

## Supplementary References

1. Ministerio de Salud del Perú. Norma técnica de salud para la vigilancia epidemiológica Y diagnóstico de laboratorio de dengue, chikungunya, zika y otras arbovirosis en el Perú. https://www.dge.gob.pe/portal/docs/tools/arbovirosis18.pdf (2017).

2. World Health Organization (WHO) & Special Programme for Research and Training in Tropical Diseases (TDR). Dengue guidelines, for diagnosis, treatment, prevention and control. (2009).

3. Normas y procedimientos para el control de las leishmaniasis en el Perú. https://www.gob.pe/institucion/minsa/informes-publicaciones/353504-normas-y-procedimientos-para-el-control-de-las-leishmaniasis-en-el-peru.

4. Ministerio de Salud del Perú. Leishmaniasis - Módulos Téchnicos. https://bvs.minsa.gob.pe/local/OGEI/795_MS-OGE106.pdf (2000).

5. Castagnetto, J. ubigeo. https://github.com/jmcastagnetto/ubigeo/?tab=readme-ov-file (2021).

6. Plataforma Nacional de Datos Abiertos. Plataforma Nacional de Datos Abiertos https://www.datosabiertos.gob.pe/dataset/establecimientos-de-salud (2024).

7. Open Spatial Demographic Data and Research. WorldPop https://www.worldpop.org/ (2024).

8. Jensen, K. E., et al. Small scale migration along the interoceanic highway in Madre de Dios, Peru: an exploration of community perceptions and dynamics due to migration. BMC Int. Health Hum. Rights 18, 12 (2018).

9. Carrasco-Escobar, G., Manrique, E., Tello-Lizarraga, K. & Miranda, J. J. Travel Time to Health Facilities as a Marker of Geographical Accessibility Across Heterogeneous Land Coverage in Peru. Front. Public Health 8, (2020).

10. Cabezas, C. et al. Dengue en el Perú: a un cuarto de siglo de su reemergencia. Rev. Peru. Med. Exp. Salud Pública 146–156 (2015) doi:10.17843/rpmesp.2015.321.1587.

11. Gutiérrez, L. A. & https://www.facebook.com/pahowho. PAHO/WHO Data - Bolivia - Casos de dengue | OPS/OMS. Pan American Health Organization / World Health Organization https://www3.paho.org/data/index.php/es/temas/indicadores-dengue/dengue-subnacional/536-bol-dengue-casos-es.html (2015).

12. (DATASUS, Ministério da Saúde do Brasil, 2022). Doenças e Agravos de Notificação – 2007 em Diante (SINAN). https://datasus.saude.gov.br/acesso-a-informacao/doencas-e-agravos-de-notificacao-de-2007-em-diante-sinan/.

13. Ministerio de Transportes y Comunicaciones. https://portal.mtc.gob.pe/estadisticas/transportes.html.

